# Efficacy, effectiveness and safety of transcranial magnetic stimulation for bipolar depression: A systematic review and meta-analysis

**DOI:** 10.1101/2025.03.19.25324258

**Authors:** Fabiana Ventura, Pedro Frias, Daniel Rodrigues da Silva, Alexander McGirr, Gonçalo Cotovio, Albino J. Oliveira-Maia

**Author notes:** Co-corresponding authors at: Gonçalo Cotovio, MD, PhD, Albino J. Oliveira-Maia, MD, MPH, PhD, Champalimaud Research and Clinical Centre, Champalimaud Centre for the Unkown, Av. Brasilia 1400-038, Lisbon, Portugal; (ext. 4065). Equally contributing first authors (Fabiana Ventura, Pedro Frias). Equally contributing senior authors (Gonçalo Cotovio, Albino J. Oliveira-Maia).

## Abstract

**Background:** Repetitive transcranial magnetic stimulation (rTMS) is a neuromodulatory treatment cleared by Food and Drug Administration (FDA) for use in major depressive disorder (MDD). Recently, FDA granted breakthrough status for rTMS as a therapeutic option for bipolar depression (BDep). Nonetheless, efficacy and safety for BDep are not yet sufficiently established and there is no guidance regarding patient and protocol selection. Here, we systematically reviewed the literature on TMS for BDep, to synthesize the evidence on efficacy, effectiveness and safety.

**Methods:** We systematically searched four literature databases for studies published between 1995-2024 treating participants with acute bipolar depression. The primary outcome for meta-analysis was change in mean depression severity scores from baseline. Determinants of treatment response were assessed using meta-regression and sub-group meta-analyses.

**Results:** Forty-nine articles were included, representing a total of 1593 patients with BDep. Active TMS had superior antidepressant efficacy relative to sham in RCTs (Cohen’s d=0.43; 95%CI 0.21-0.64; p<0.001; N=18). Rates of treatment-emergent mania or hypomania were low and equivalent to those found for sham (OR=1.3; 95%CI 0.7-2.5). A large effect size for antidepressant effectiveness was found when pooling active arms of RCTs with data from non-controlled studies (Cohen’s d=1.40 95%CI 1.2-1.6; p<0.001; N=39), with rates of response (47.2%, 95%CI 35.3-59.1) and remission (27.1%, 95%CI 22.1-32.2) similar to those described for MDD, and preserved in sub-analyses for high frequency protocols, including iTBS, delivered to the left DLPFC, and low frequency protocols delivered to right DLPFC. Baseline illness severity, shorter illness duration and longer treatment courses were predictors of greater antidepressant effect.

**Conclusions:** TMS is efficacious and safe in BDep, with response and remission rates on par with unipolar depression. Longer protocols, namely with high-frequency or iTBS to the L-DLPFC and low-frequency to the R-DLPFC, have the best results, and patient selection may further improve clinical outcomes.

## 1. INTRODUCTION

The global lifetime prevalence of bipolar disorder (BD) is estimated to be 2% (1–3) with depressive episodes representing 70-80% of symptomatic periods and contributing most strongly to disability(4, 5). Unfortunately, there are few pharmacological treatments approved for bipolar depression(6), with many patients failing to respond to available strategies and/or suffering from intolerable side effects, which further limits the available treatment options for this condition. Repetitive transcranial magnetic stimulation (rTMS) is a non-invasive neuromodulation technique, cleared by the Food and Drug Administration (FDA) for episodes of major depression (7). The treatment works by modulating the activity of target brain regions using electromagnetic pulses, delivered by a coil placed over the patient’s scalp(8). Different TMS protocols may facilitate or inhibit neuronal activity in the targeted brain region. High frequency protocols, such as >5Hz standard rTMS or intermittent theta burst stimulation (iTBS; 3-pulse 50Hz bursts, delivered at 5Hz) are typically facilitatory, while low frequency (1Hz) standard rTMS and continuous TBS protocols (cTBS) are typically inhibitory(9).

Treatment with rTMS has been described to be better tolerated when compared to other methods for major depression(10), since it is devoid of drug interactions and side effects commonly observed with pharmacological treatment strategies, such as antidepressants, mood-stabilizers and antipsychotics(11). Given this well-established safety and tolerability profile, as well as the lack of alternatives, rTMS has been explored as a treatment option also for bipolar depression (BDep), and was granted breakthrough status to treat bipolar depression (BDep) by the FDA(12). However, there is controversy regarding the role of TMS in the management of BDep. Indeed, available sham-controlled trials testing the efficacy of TMS for BDep have had both positive (13, 14) and negative (15) results.

Meta-analyses of the trials testing TMS for BDep support superiority over sham(16–18). Real-world data is also indicative of effectiveness, with conflicting reports of whether it is superior (19) or inferior (20) to the effectiveness in episodes of major depression. However, previous reviews and meta-analyses included a preponderance of studies with small or very small sample sizes, with substantial variability in the antidepressant effects of TMS across studies. Importantly, this has not allowed for analyses to inform on determinants of these effects, that could guide selection of the most appropriate treatment protocols or patients. Here, we conducted an updated systematic review to synthesize the evidence for TMS in BDep, using randomized controlled trials to assess efficacy and safety, and both controlled and uncontrolled trials for assessment of effectiveness. With the data for effectiveness, we further tested the TMS protocols and parameters, as well as patient characteristics, associated with greater response to treatment.

## 2. METHODS AND MATERIALS

### 2.1. Protocol and registration

The study methodology was designed according to Cochrane recommendations (21) and PRISMA guidelines (22) and published *a priori* in a written protocol in Prospero (registration number PROSPERO: 2022 CRD42022330838) available online (https://www.crd.york.ac.uk/prospero/display_record.php?RecordID=330838).

### 2.2. Information sources and search strategy

The systematic literature review included randomized, sham-controlled trials (RCTs) assessing the efficacy of TMS in patients with BDep, specifically on depression severity rating scores and treatment response and/or remission rates, as defined per study. For meta-regression analyses, only active arms of RCTs were considered, and open label trials and retrospective studies were also included. Case reports, case series and animal studies were excluded. Studies were identified through electronic searches in MEDLINE/PubMed, Web of Science, The Cochrane Library and Embase databases. The search included publications from 1^st^ January 1995 to 24 July 2024, using the following syntax: (transcranial stimulation OR TMS OR transcranial magnetic stimulation OR non-invasive brain stimulation OR NIBS OR intermittent theta-burst stimulation OR iTBS OR theta-burst stimulation OR TBS OR continuous theta-burst stimulation OR cTBS OR deep TMS OR dTMS OR repetitive transcranial magnetic stimulation OR rTMS) AND (depressive disorder OR resistant depression OR depressive episode OR depression) AND (bipolar OR bipolar disorder OR manic depressive). Filters were applied to restrict search results to adult human subjects (please see Table S1 for further details). Additionally, we searched references lists of the initially selected studies, as well as of systematic reviews and meta-analyses, for additional eligible articles. Only articles in English, Spanish, French, Portuguese, or Mandarin were considered.

### 2.3. Study selection and eligibility criteria

The studies identified in the literature search were independently selected by two researchers (FV and PF) in sequential phases of title, abstract and full-text review, with consensus at the end of each step and disagreements resolved by a third researcher (GC). Only studies with participants older than 18 years old with BD and an ongoing episode of depression, as defined by the Diagnostic and Statistical Manual of Mental Disorders (DSM-III or later edition), or its equivalent in the International Classification of Diseases (ICD-9 or later editions), were included. Studies were excluded if they did not provide efficacy data regarding depression severity scores (mean and standard deviation [SD] or standard error [SE]) nor data regarding treatment response and/or remission rates, and such data was not provided upon request to authors (Table S2). Studies reporting data from patients diagnosed with either bipolar or unipolar depression were included only if BDep data could be isolated, either from the article, or as provided by request to the author (Table S2).

### 2.4. Data extraction, data items and risk of bias

Data was independently extracted by two researchers (FV and PF) with disagreements resolved by a third researcher (GC). For each paper, the following information was collected: first author name, title, journal, publication year, study type, total sample size, treatment groups (e.g., active vs. sham). For each treatment group the following data was extracted: sample size, mean age, percentage of female participants, mean duration of illness (years), mean number of depressive episodes, mean duration of current depressive episode (months), use of medication, symptom severity before and after treatment (Hamilton Depression Rating Scale, HDRS; Montgomery-Asberg Depression Scale, MADRS; Beck Depression Inventory, BDI), TRD criteria. Parameters for TMS stimulation protocols were also extracted: stimulator type, coil, method of determination of motor threshold, intensity (% of resting motor threshold), stimulation target (site, method of determination), frequency (Hz), trains (N), inter-train interval (s), pulses per session (N), total sessions (N), sessions/day (N). Reported side effects were also extracted, with a specific focus on treatment emergent mania. Lack of report of a side effect was considered as evidence that it did not occur, to allow for meta-analysis. Study authors were contacted to acquire or clarify data not reported in the available articles (Table S2), including clinical and demographic data (Table S3). Study quality was defined by consensus between two researchers (FV and PF) according to the Cochrane risk-of-bias tool (RoB2) for randomized trials (23) and Newcastle-Ottawa Quality Assessment Scale for cohort studies(24). Each scale was divided into 3 levels (level 1 – high quality, level 2 – moderate quality, level 3 – low quality) to compute a single quality-related variable to test in meta-regression analyses (details below).

### 2.5. Statistical analysis

Analyses were performed using StataCorp. 2017 (Stata Statistical Software: Release 15; StataCorp LP, College Station, TX). Two levels of data analysis were considered, namely RCTs for meta-analyses to determine treatment efficacy, and uncontrolled studies pooled with the active arms of controlled studies for meta-analyses and meta-regressions to determine treatment effectiveness and determinants of antidepressant effect. Open-label extensions of RCTs were not considered in analyses of uncontrolled data due to potential error in determining the effect across sequential TMS protocols. Hence, we opted to always use the available data only for the first TMS treatment in both RCT and open label studies.

Measures of treatment efficacy were analyzed for meta-analysis, with the primary outcome defined as the difference between sham and active TMS in the mean change of depression severity after treatment (continuous outcome). The difference between sham and active TMS in response or remission rates (categorical outcomes), were defined as secondary outcomes. The mean difference in depression severity before and after active rTMS as well as response and remission rates were the measures of treatment effectiveness considered for both meta-analyses and meta-regressions. The standard deviation for the mean difference in depression severity (SDMD) was calculated using the following formula:

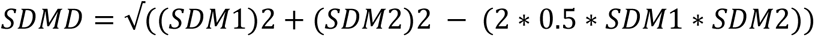

where SDM1 and SDM2 are the pre- and post-treatment Standard Deviation of the Mean depression severity, respectively. The *r* coefficient was set at 0.5 by consensus and according to previous reports(25). When the 95% Confidence Interval (CI) was available instead of SD, we used the following formula to calculate the latter:

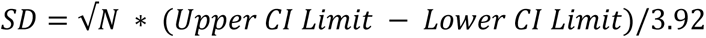

where N is total sample size(20). When the SD of the parameters of interest was not reported, the average SD value of all studies was used(26). Several methods were used to account for potential mathematical constraints. When SDMD was not possible to calculate, we used the average SDMD in the remaining studies, according to previous practice(27). When considering response or remission rates, to allow CI calculation for 0 and 100%, 0.1% and 99.9% were used instead, respectively. In studies containing zero observed events, cell count was provided according to Treatment Arm Continuity Correction (TA-CC), as performed by others(28). TMS Sensitivity analyses of meta-analytic results were performed excluding studies for which these strategies were necessary.

Cohen’s d was used as a measure of effect size (ES) of outcomes, given the range in sample sizes across studies. For categorical outcomes, odds ratios (OR) were used to determine effect size. Number needed to treat (NNT) for efficacy and number need to harm (NNH) for side-effects were computed where appropriate.

All metanalyses were performed using random-effect models(29), since we found high heterogeneity (I^2^>35% and χ^2^ test p<0.1) between studies(21). When comparing sham to active TMS, we used funnel plots and Egger’s regression tests to assess for potential publication bias, and Duval and Tweedie’s trim-and-fill method to account for unpublished studies within the field, providing adjusted summary effects. To identify variables that could determine or predict treatment effectiveness in active arms of sham-controlled trials and in non-controlled studies, we conducted subgroup metanalyses as well as meta-regressions. The former were conducted for categorical variables while the latter were used for continuous variables. Meta-regressions were only performed if 10 or more studies were available for the tested variable, according to best practice(21).

## 3. RESULTS

### 3.1. Literature review and Studies Synthesis

The initial literature search yielded 1058 articles, after removing duplicates. After title, abstract and full-text review (Fig. 1), 49 articles were eligible, 29 of which were RCTs (13–15, 30–55), 12 were open-label clinical trials (56–67) and 8 were retrospective studies (19, 20, 68–73). A total of 1593 patients diagnosed with BDep were included in these studies (351 bipolar disorder type I, 388 bipolar disorder type II and 854 not classified/mixed sample). Of these, 1060 patients were treated with active TMS and 334 patients with sham TMS. In the active group, mean age at baseline was 44.4±7.8 years-old, mean duration of current episode was 8.4±5.96 months, mean number of depressive episodes was 5.2±3.5 and mean illness duration was 17.2±5.96 years. In the sham group, mean age at baseline was 42.3±8.98 years, mean length of current episode was 8.7±5.8 months, mean number of depressive episodes was 6.1±4.3 and mean duration of illness was 17.5±3.8 years (Table 1). Among the 49 eligible studies, 27 also reported data from patients with unipolar depression (19, 20, 37–50, 56–61, 68–71, 74). Twenty-five studies included patients diagnosed with treatment resistant BDep, and 38 reported concomitant medication (Table S4). Different TMS protocols were used across studies (Table S5). Risk of bias was moderate to low, and study quality was moderate to high (Tables S6 and S7).

**Figure 1.**
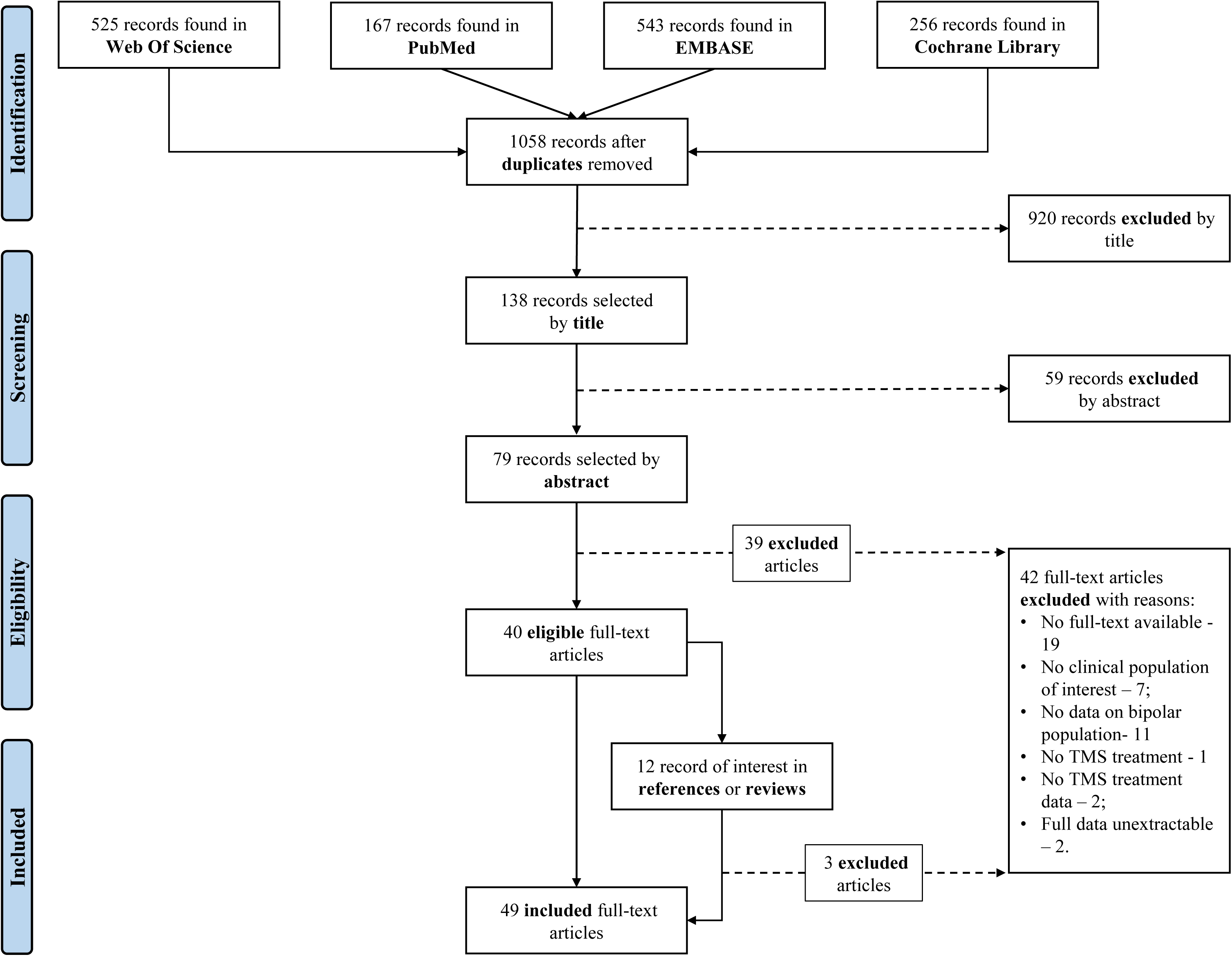
Article selection flowchart. A systematic review was performed according to PRISMA Guidelines (22) and, from an initial pool of 1058 articles, after removing duplicates, 49 were included: 29 randomized-controlled trials, 12 open-label clinical trials and 8 retrospective studies.

**Table 1.**
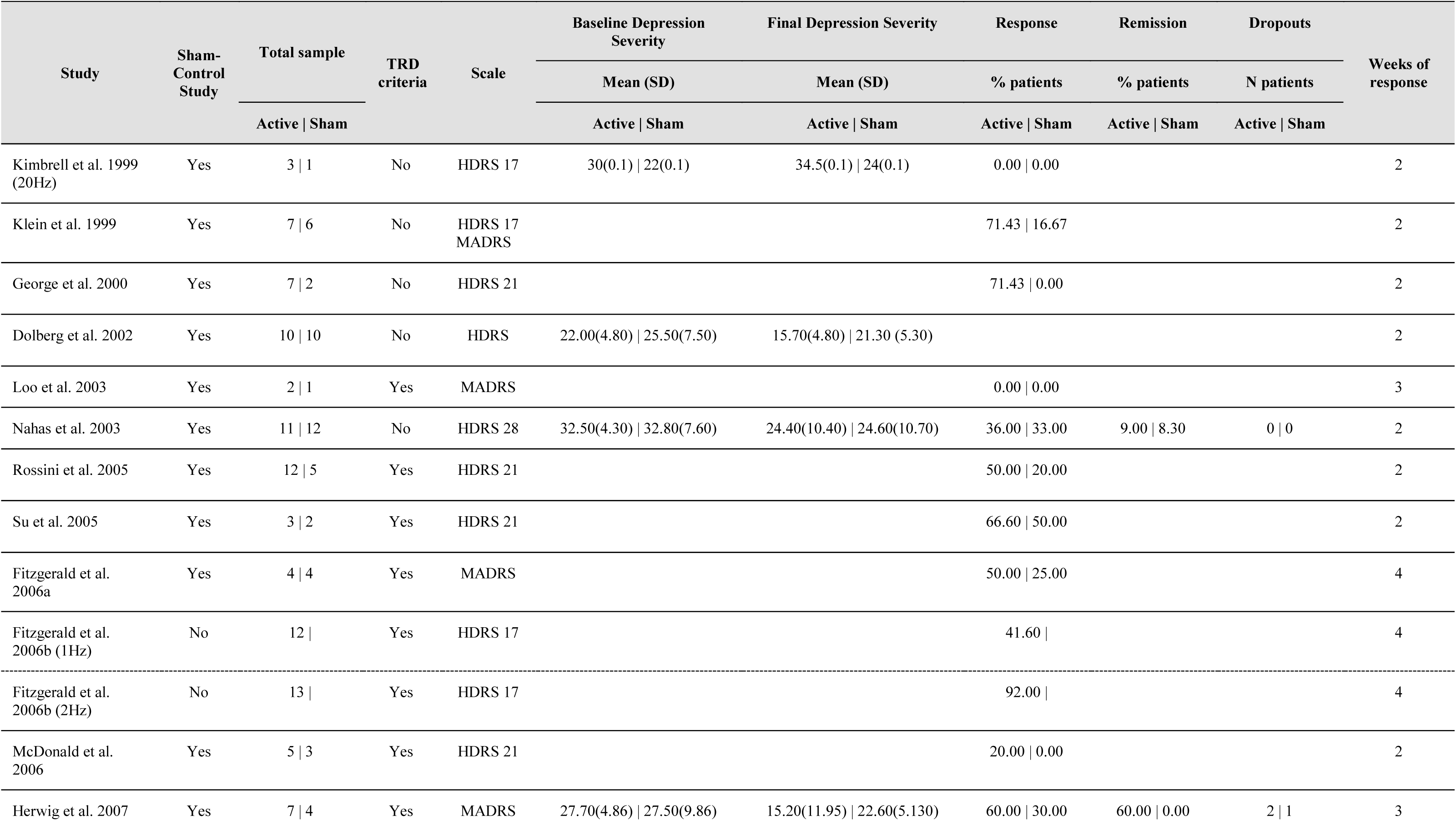
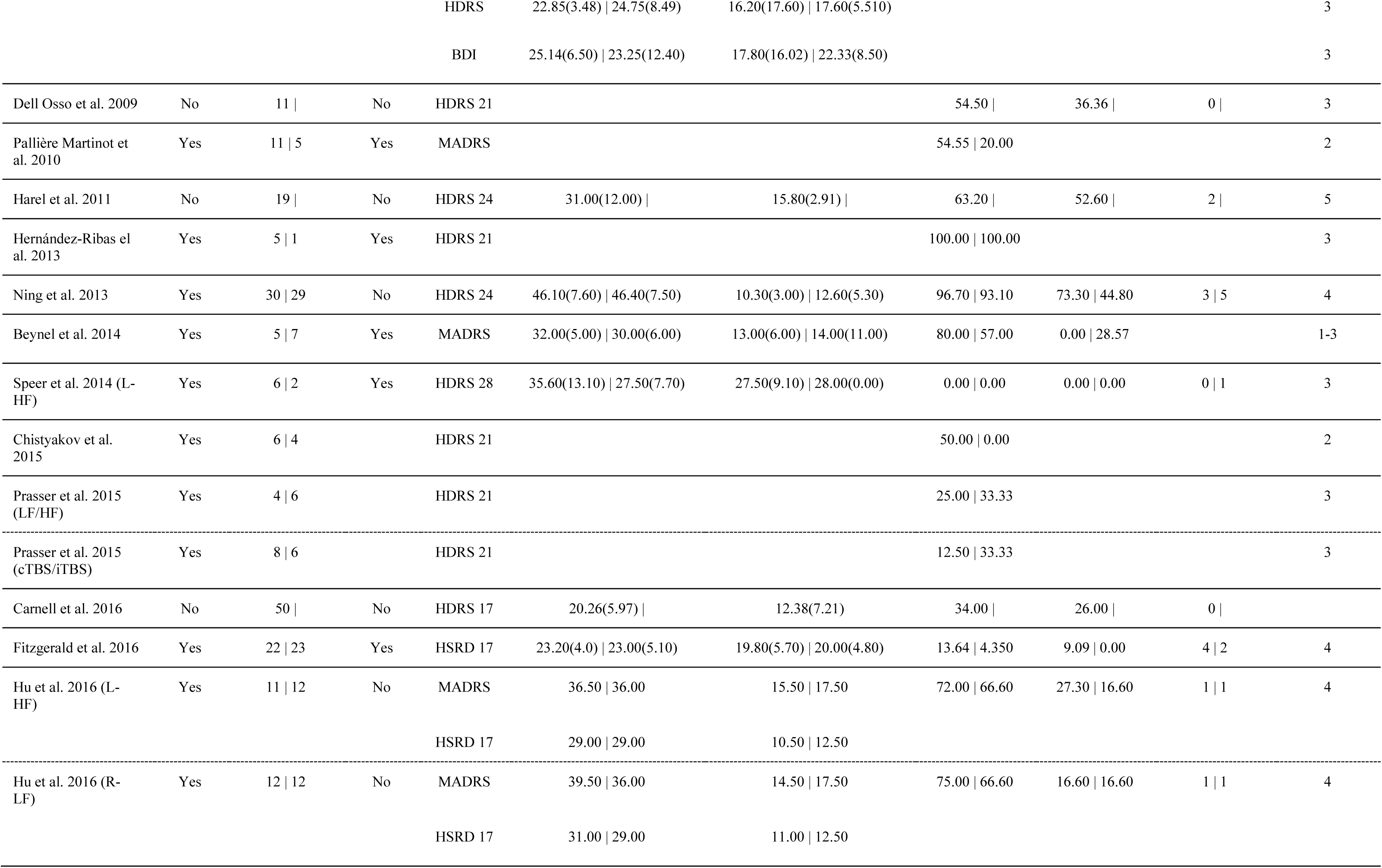
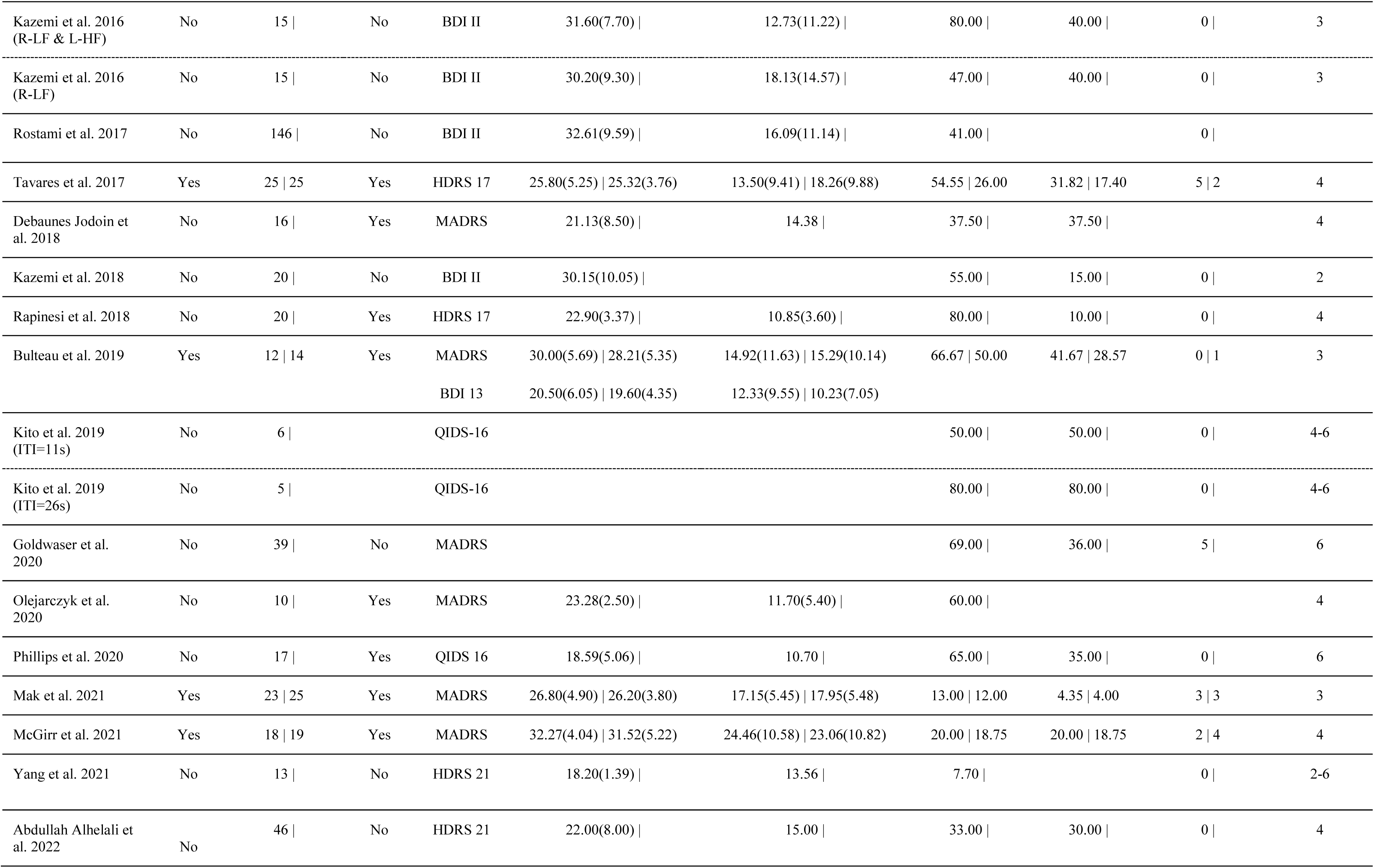
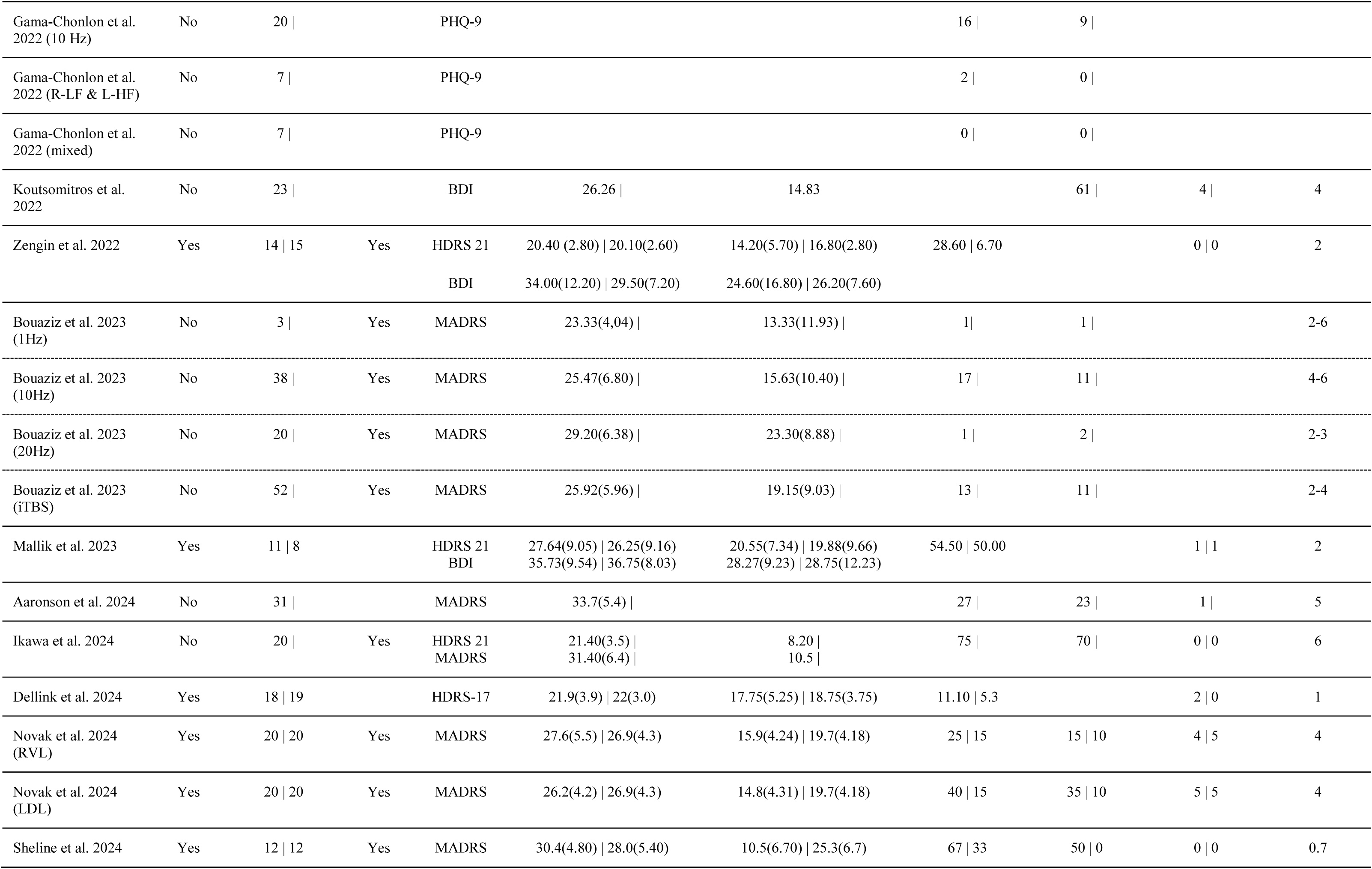
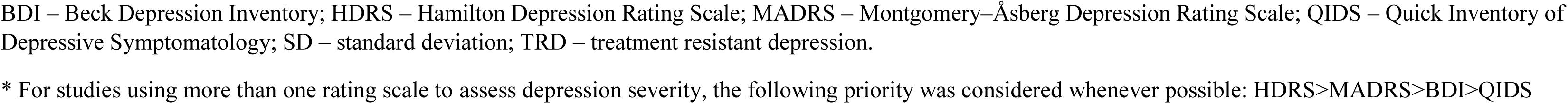
Summary table for the eligible studies.

Table References 1. Alhelali A, Almheiri E, Abdelnaim M, Weber FC, Langguth B, Schecklmann M, et al. Effectiveness of Repetitive Transcranial Magnetic Stimulation in the Treatment of Bipolar Disorder in Comparison to the Treatment of Unipolar Depression in a Naturalistic Setting. Brain Sci. 2022;12(3). 2. Beynel L, Chauvin A, Guyader N, Harquel S, Szekely D, Bougerol T, et al. What saccadic eye movements tell us about TMS-induced neuromodulation of the DLPFC and mood changes: a pilot study in bipolar disorders. Front Integr Neurosci. 2014;8:65. 3. Bulteau S, Beynel L, Marendaz C, Dall’Igna G, Peré M, Harquel S, et al. Twice-daily neuronavigated intermittent theta burst stimulation for bipolar depression: A Randomized Sham-Controlled Pilot Study. Neurophysiol Clin. 2019;49(5):371-5. 4. Carnell BL, Clarke P, Gill S, Galletly CA. How effective is repetitive transcranial magnetic stimulation for bipolar depression? J Affect Disord. 2017;209:270-2. 5. Dell’Osso B, Mundo E, D’Urso N, Pozzoli S, Buoli M, Ciabatti M, et al. Augmentative repetitive navigated transcranial magnetic stimulation (rTMS) in drug-resistant bipolar depression. Bipolar Disord. 2009;11(1):76-81. 6. Desbeaumes Jodoin V, Miron JP, Lespérance P. Safety and Efficacy of Accelerated Repetitive Transcranial Magnetic Stimulation Protocol in Elderly Depressed Unipolar and Bipolar Patients. Am J Geriatr Psychiatry. 2019;27(5):548-58. 7. Dolberg OT, Dannon PN, Schreiber S, Grunhaus L. Transcranial magnetic stimulation in patients with bipolar depression: a double blind, controlled study. Bipolar Disord. 2002;4 Suppl 1:94-5. 8. Fitzgerald PB, Benitez J, de Castella A, Daskalakis ZJ, Brown TL, Kulkarni J. A randomized, controlled trial of sequential bilateral repetitive transcranial magnetic stimulation for treatment-resistant depression. Am J Psychiatry. 2006;163(1):88-94. 9. Fitzgerald PB, Hoy KE, Elliot D, McQueen S, Wambeek LE, Daskalakis ZJ. A negative double-blind controlled trial of sequential bilateral rTMS in the treatment of bipolar depression. J Affect Disord. 2016;198:158-62. 10. Fitzgerald PB, Huntsman S, Gunewardene R, Kulkarni J, Daskalakis ZJ. A randomized trial of low-frequency right-prefrontal-cortex transcranial magnetic stimulation as augmentation in treatment-resistant major depression. Int J Neuropsychopharmacol. 2006;9(6):655-66. 11. Goldwaser EL, Daddario K, Aaronson ST. A retrospective analysis of bipolar depression treated with transcranial magnetic stimulation. Brain Behav. 2020;10(12):e01805. 12. Harel EV, Zangen A, Roth Y, Reti I, Braw Y, Levkovitz Y. H-coil repetitive transcranial magnetic stimulation for the treatment of bipolar depression: an add-on, safety and feasibility study. World J Biol Psychiatry. 2011;12(2):119-26. 13. Herwig U, Fallgatter AJ, Höppner J, Eschweiler GW, Kron M, Hajak G, et al. Antidepressant effects of augmentative transcranial magnetic stimulation: randomised multicentre trial. Br J Psychiatry. 2007;191:441-8. 14. Hu SH, Lai JB, Xu DR, Qi HL, Peterson BS, Bao AM, et al. Efficacy of repetitive transcranial magnetic stimulation with quetiapine in treating bipolar II depression: a randomized, double-blinded, control study. Sci Rep. 2016;6:30537. 15. Kazemi R, Rostami R, Khomami S, Baghdadi G, Rezaei M, Hata M, et al. Bilateral Transcranial Magnetic Stimulation on DLPFC Changes Resting State Networks and Cognitive Function in Patients With Bipolar Depression. Front Hum Neurosci. 2018;12:356. 16. Kazemi R, Rostami R, Khomami S, Horacek J, Brunovsky M, Novak T, et al. Electrophysiological correlates of bilateral and unilateral repetitive transcranial magnetic stimulation in patients with bipolar depression. Psychiatry Res. 2016;240:364-75. 17. Kito S, Miyazi M, Nakatani H, Matsuda Y, Yamazaki R, Okamoto T, et al. Effectiveness of high-frequency left prefrontal repetitive transcranial magnetic stimulation in patients with treatment-resistant depression: A randomized clinical trial of 37.5-minute vs 18.75-minute protocol. Neuropsychopharmacol Rep. 2019;39(3):203-8. 18. Mak ADP, Neggers SFW, Leung ONW, Chu WCW, Ho JYM, Chou IWY, et al. Antidepressant efficacy of low-frequency repetitive transcranial magnetic stimulation in antidepressant-nonresponding bipolar depression: a single-blind randomized sham-controlled trial. Int J Bipolar Disord. 2021;9(1):40. 19. McGirr A, Vila-Rodriguez F, Cole J, Torres IJ, Arumugham SS, Keramatian K, et al. Efficacy of Active vs Sham Intermittent Theta Burst Transcranial Magnetic Stimulation for Patients With Bipolar Depression: A Randomized Clinical Trial. JAMA Netw Open. 2021;4(3):e210963. 20. Nahas Z, Kozel FA, Li X, Anderson B, George MS. Left prefrontal transcranial magnetic stimulation (TMS) treatment of depression in bipolar affective disorder: a pilot study of acute safety and efficacy. Bipolar Disord. 2003;5(1):40-7. 21. Olejarczyk E, Zuchowicz U, Wozniak-Kwasniewska A, Kaminski M, Szekely D, David O. The Impact of Repetitive Transcranial Magnetic Stimulation on Functional Connectivity in Major Depressive Disorder and Bipolar Disorder Evaluated by Directed Transfer Function and Indices Based on Graph Theory. Int J Neural Syst. 2020;30(4):2050015. 22. Phillips AL, Burr RL, Dunner DL. Repetitive Transcranial Magnetic Stimulation in the Treatment of Bipolar Depression: Experience From a Clinical Setting. J Psychiatr Pract. 2020;26(1):37-45. 23. Rapinesi C, Kotzalidis GD, Ferracuti S, Girardi N, Zangen A, Sani G, et al. Add-on high frequency deep transcranial magnetic stimulation (dTMS) to bilateral prefrontal cortex in depressive episodes of patients with major depressive disorder, bipolar disorder I, and major depressive with alcohol use disorders. Neurosci Lett. 2018;671:128-32. 24. Rostami R, Kazemi R, Nitsche MA, Gholipour F, Salehinejad MA. Clinical and demographic predictors of response to rTMS treatment in unipolar and bipolar depressive disorders. Clin Neurophysiol. 2017;128(10):1961-70. 25. Speer AM, Wassermann EM, Benson BE, Herscovitch P, Post RM. Antidepressant efficacy of high and low frequency rTMS at 110% of motor threshold versus sham stimulation over left prefrontal cortex. Brain Stimul. 2014;7(1):36-41. 26. Tavares DF, Myczkowski ML, Alberto RL, Valiengo L, Rios RM, Gordon P, et al. Treatment of Bipolar Depression with Deep TMS: Results from a Double-Blind, Randomized, Parallel Group, Sham-Controlled Clinical Trial. Neuropsychopharmacology. 2017;42(13):2593-601. 27. Yang YB, Chan P, Rayani K, McGirr A. Comparative Effectiveness of Repetitive Transcranial Magnetic Stimulation in Unipolar and Bipolar Depression. Can J Psychiatry. 66. United States2021. p. 313-5. 28. Zengin G, Topak OZ, Atesci O, Culha Atesci F. The Efficacy and Safety of Transcranial Magnetic Stimulation in Treatment-Resistant Bipolar Depression. Psychiatr Danub. 2022;34(2):236-44. 29. Ning L, Xue-Yi W, Zhen-Zhou Q, Mei S. A 4-week single-blind randomized controlled trial of repetitive transcranial magnetic stimulation combined with lithium and quetiapine in treatment of patients with bipolar depression. Chinese mental health journal. 2013;27(12):896-900. 30. Klein E, Kreinin I, Chistyakov A, Koren D, Mecz L, Marmur S, et al. Therapeutic efficacy of right prefrontal slow repetitive transcranial magnetic stimulation in major depression: a double-blind controlled study. Arch Gen Psychiatry. 1999;56(4):315-20. 31. Kimbrell TA, Little JT, Dunn RT, Frye MA, Greenberg BD, Wassermann EM, et al. Frequency dependence of antidepressant response to left prefrontal repetitive transcranial magnetic stimulation (rTMS) as a function of baseline cerebral glucose metabolism. Biol Psychiatry. 1999;46(12):1603-13. 32. George MS, Nahas Z, Molloy M, Speer AM, Oliver NC, Li XB, et al. A controlled trial of daily left prefrontal cortex TMS for treating depression. Biol Psychiatry. 2000;48(10):962-70. 33. Loo CK, Mitchell PB, Croker VM, Malhi GS, Wen W, Gandevia SC, et al. Double-blind controlled investigation of bilateral prefrontal transcranial magnetic stimulation for the treatment of resistant major depression. Psychol Med. 2003;33(1):33-40. 34. Rossini D, Lucca A, Zanardi R, Magri L, Smeraldi E. Transcranial magnetic stimulation in treatment-resistant depressed patients: a double-blind, placebo-controlled trial. Psychiatry Res. 2005;137(1-2):1-10. 35. Su TP, Huang CC, Wei IH. Add-on rTMS for medication-resistant depression: a randomized, double-blind, sham-controlled trial in Chinese patients. J Clin Psychiatry. 2005;66(7):930-7. 36. McDonald WM, Easley K, Byrd EH, Holtzheimer P, Tuohy S, Woodard JL, et al. Combination rapid transcranial magnetic stimulation in treatment refractory depression. Neuropsychiatr Dis Treat. 2006;2(1):85-94. 37. Paillère Martinot ML, Galinowski A, Ringuenet D, Gallarda T, Lefaucheur JP, Bellivier F, et al. Influence of prefrontal target region on the efficacy of repetitive transcranial magnetic stimulation in patients with medication-resistant depression: a [(18)F]-fluorodeoxyglucose PET and MRI study. Int J Neuropsychopharmacol. 2010;13(1):45-59. 38. Hernández-Ribas R, Deus J, Pujol J, Segalàs C, Vallejo J, Menchón JM, et al. Identifying brain imaging correlates of clinical response to repetitive transcranial magnetic stimulation (rTMS) in major depression. Brain Stimul. 2013;6(1):54-61. 39. Chistyakov AV, Kreinin B, Marmor S, Kaplan B, Khatib A, Darawsheh N, et al. Preliminary assessment of the therapeutic efficacy of continuous theta-burst magnetic stimulation (cTBS) in major depression: a double-blind sham-controlled study. J Affect Disord. 2015;170:225-9. 40. Prasser J, Schecklmann M, Poeppl TB, Frank E, Kreuzer PM, Hajak G, et al. Bilateral prefrontal rTMS and theta burst TMS as an add-on treatment for depression: a randomized placebo controlled trial. World J Biol Psychiatry. 2015;16(1):57-65. 41. Koutsomitros T, van der Zee KT, Evagorou O, Schuhmann T, Zamar AC, Sack AT. A Different rTMS Protocol for a Different Type of Depression: 20.000 rTMS Pulses for the Treatment of Bipolar Depression Type II. Journal of Clinical Medicine. 2022;11(18). 42. Bouaziz N, Laidi C, Bulteau S, Berjamin C, Thoms F, Moulier V, et al. Real world transcranial magnetic stimulation for major depression: a multisite naturalistic, retrospective study. Brain Stimulation. 2023;16(1):384-5. 43. Mallik G, Mishra P, Garg S, Dhyani M, Tikka SK, Tyagi P. Safety and Efficacy of Continuous Theta Burst “Intensive” Stimulation in Acute-Phase Bipolar Depression A Pilot, Exploratory Study. Journal of Ect. 2023;39(1):28-33. 44. Ikawa H, Osawa R, Takeda Y, Sato A, Mizuno H, Noda Y. Real-world retrospective study of repetitive transcranial magnetic stimulation (TMS) treatment for bipolar and unipolar depression using TMS registry data in Tokyo. Heliyon. 2024;10(5):e27288. 45. Gama-Chonlon L, Scanlan JM, Allen RM. Could bipolar depressed patients respond better to rTMS than unipolar depressed patients? A naturalistic, observational study. Psychiatry Research. 2022;312. 46. Aaronson ST, Goldwaser EL, Croarkin PE, Geske JR, LeMahieu A, Sklar JH, et al. A Pilot Study of High-Frequency Transcranial Magnetic Stimulation for Bipolar Depression. J Clin Psychiatry. 2024;85(2). 47. Sheline YI, Makhoul W, Batzdorf AS, Nitchie FJ, Lynch KG, Cash R, et al. Accelerated Intermittent Theta-Burst Stimulation and Treatment-Refractory Bipolar Depression: A Randomized Clinical Trial. JAMA Psychiatry. 2024. 48. Novák T, Kostýlková L, Bareš M, Renková V, Hejzlar M, Renka J, et al. Right ventrolateral and left dorsolateral 10 Hz transcranial magnetic stimulation as an add-on treatment for bipolar I and II depression: a double-blind, randomised, three-arm, sham-controlled study. World J Biol Psychiatry. 2024;25(5):304-16. 49. Dellink A, Hebbrecht K, Zeeuws D, Baeken C, De Fré G, Bervoets C, et al. Continuous theta burst stimulation for bipolar depression: A multicenter, double-blind randomized controlled study exploring treatment efficacy and predictive potential of kynurenine metabolites. J Affect Disord. 2024;361:693-701.

### 3.2. Treatment Efficacy & Tolerability

When comparing sham to active TMS, we found a significant effect favoring active TMS (Cohen’s d=0.43; 95% CI 0.21-0.64; p<0.001; N=18; Figure 2a). Overall OR for response was 2.2 (95% CI 1.5-3.3; p<0.001; N=28; NNT=7; figure 2b) and for remission 2.3 (95% CI 1.4-3.9; p=0.001; N=16; NNT=8; figure 2c). Findings were consistent in the sensitivity analyses excluding studies where we used the strategies described in Methods to account for potential mathematical constraints (data not shown). Dropout rates were similar between active and sham interventions (OR=0.9; 95% CI 0.5-1.6; p=0.8; N=16; figure 2d). The number of available studies allowing for assessment of publication bias was 49 (75), and visual inspection of funnel plots suggested potential publication bias (Figure S1). However, Eggers’s tests supported the presence of publication bias only for the response rate metanalysis (p=0.04), but not the metanalyses for depression score improvement (p=0.3), remission rate (p=0.3) and dropout rate (p=0.6). Importantly, the Trim-Fill Duval-Tweedie Test yielded unchanged effects sizes for the depression score (Cohen’s d=0.43; 95% CI 0.21-0.64) and dropout rate metanalyses (OR=1.1; 95% CI 0.6-1.7), and increased ORs for response rate (3.4; 95% CI 2.4-4.5) and remission rate (4.2; 95% CI 2.0-6.3), supporting that results are robust to publication bias. Finally, leave one out metanalyses confirmed that no single study drove the results.

**Figure 2.**
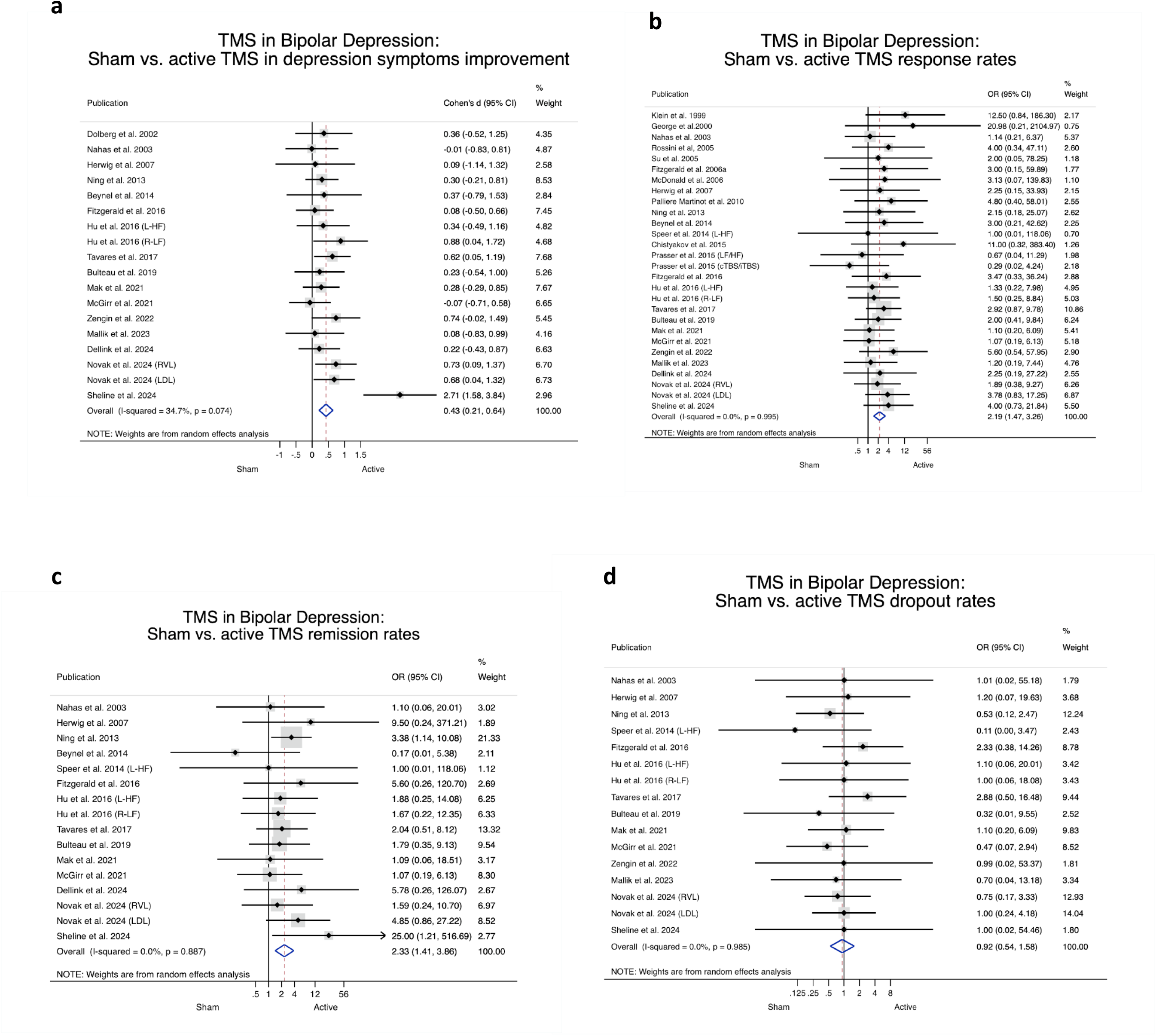
Forest plots of random effects meta-analyses. There is a significant effect of TMS on depressive symptoms severity when compared to sham (a). Response and remission rates favor active treatment over sham (b, c). Dropout rates were similar between active and sham (d).

Concerning side effects, treatment-emergent hypomania/mania was reported in sixteen patients, 9 in the active group (15, 51, 52) and 5 in the sham group(51, 52), resulting in a non-significant difference between groups (OR=1.3; 95% CI 0.7-2.5; p=0.5; N=29; NNH=97; Figure S2). Only one study reported a TMS-induced generalized seizure, that was short and uncomplicated (62). Other side effects, such as mild headaches, local sensitivity, fatigue, sleepiness, and insomnia, were mild and more frequently described (13, 34, 36–38, 42–44, 50, 53, 61, 63, 69, 70, 74, 76).

### 3.3. Treatment Effectiveness

Given the evidence to support efficacy and tolerability of rTMS for BDep, we further aimed to summarize available data to inform real-world use of TMS in BDep. Hence, we performed metanalyses pooling data from active arms of sham-controlled trials with data from non-controlled studies. For analysis purposes, studies were divided into more than one active arm in papers comparing effects of distinct rTMS protocols namely Fitzgerald et al. (2006b), assessing a 1Hz and a 2Hz rTMS protocols over the left DLPFC, Speer et al. (2014), evaluating 20Hz or 1Hz protocols over the left DLPFC, Hu et al. (2016), testing 10Hz rTMS to the left DLPFC or 1Hz rTMS to the right DLPFC, Kazemi et al. (2016), comparing bilateral rTMS to 1Hz over the right DLPFC, Kito et al. (2019), comparing two left DLPFC protocols with different inter-train intervals (26s vs 11s), Gama-Chonlon et al. (2022), testing high-frequency (HF) left-side stimulation, bilateral stimulation, and a mixed protocol, Bouaziz et al. (2023), comparing 1Hz stimulation of the right DLPFC and 10Hz, 20Hz or 50Hz stimulation of the left-DLPFC, and Novak et al. (2024), testing 10 Hz rTMS applied to right ventrolateral (RVL) or left dorsolateral (LDL) prefrontal cortex. Across studies and study arms, TMS had an overall significant effect on reduction of depression symptom severity (Cohen’s d=1.4 95% CI 1.2-1.6; p<0.001; N=39; figure 3a). Mean rates of treatment response were 47.2% (95% CI 35.3-59.1; p<0.001; N=58) and of remission 27.1% (95% CI 22.1-32.2; p<0.001; N=40; Figure 3b-c). Additional analyses were performed to explore the impact of specific TMS protocol types on each of these 3 treatment outcomes. Left-sided HF-rTMS and right-sided LF-rTMS, both with 10 or more studies, were significantly associated with antidepressant effects across outcomes. Despite fewer studies available, left-sided iTBS, as well as protocols with right LF-rTMS followed by left HF-rTMS, also showed significant treatment effect across outcomes. The remaining protocols did not provide sufficient evidence and/or consistent results across outcomes (Table 2).

**Figure 3.**
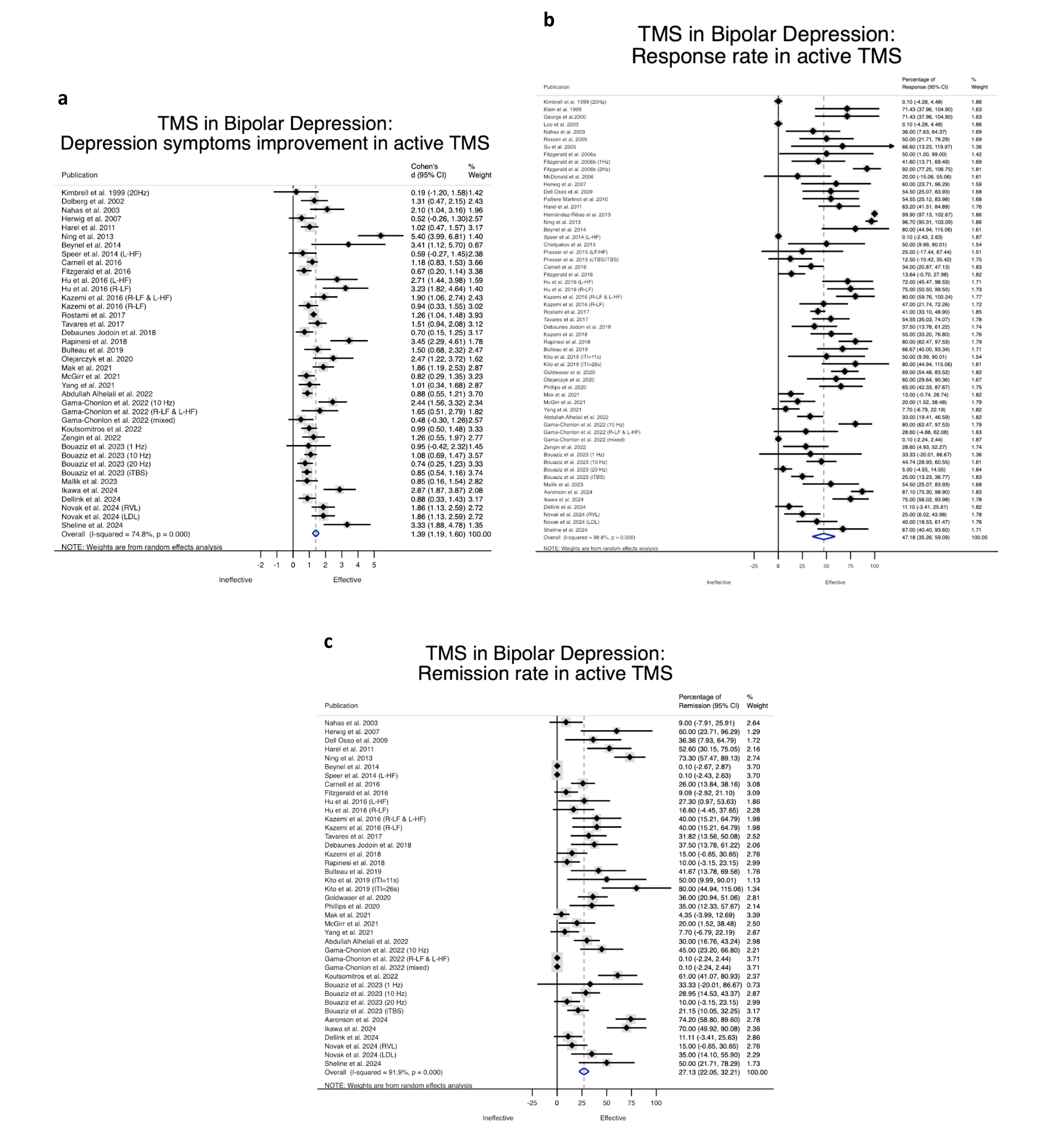
Forest plots of random effects meta-analyses. There is a significant improvement in depression symptoms with active TMS (a). Mean rate of response and remission were 47.18% (b) and 27.13% (c), respectively.

**Table 2.** Sub-group meta-analysis for different TMS protocols.

| TMS Protocols |  | Response rate |  |  | Remission rate |  |  | Dep. improvement |  |  |
| --- | --- | --- | --- | --- | --- | --- | --- | --- | --- | --- |
|  |  | N | % | p | N | % | p | N | d | p |
| Left | iTBS-L | 5 | 48.63 | <0.001 | 7 | 39.26 | 0.01 | NA | NA | NA |
|  | HF-rTMS-L | 21 | 49.95 | <0.001 | 7 | 38.14 | <0.001 | 15 | 1.19 | <0.001 |
| Right | HF-rTMS-R | 1 | 25.00 | 0.01 | 1 | 15.00 | 0.060 | 1 | 1.86 | <0.001 |
|  | LF-rTMS-R | 10 | 61.15 | <0.001 | 5 | 23.29 | 0.004 | 6 | 2.47 | <0.001 |
|  | cTBS-R | 3 | 35.60 | 0.03 | 1 | 11.11 | 0.13 | 2 | 0.87 | <0.001 |
| Bilateral | HF-rTMS-BL | 2 | 39.58 | 0.32 | 1 | 10.00 | 0.14 | 1 | 3.45 | <0.001 |
|  | LF-rTMS-R followed by HF-rTMS-L | 5 | 45.06 | 0.004 | 3 | 18.10 | 0.02 | 2 | 1.23 | 0.04 |
|  | cTBS-R followed by iTBS-L | 1 | 12.50 | 0.29 | NA | NA | NA | NA | NA | NA |

BL – Bilateral; Dep. – Depression; cTBS – continuous Theta Burst Stimulation; HF – High frequency; iTBS – intermittent Theta Burst Stimulation; L – Left; LF – Low Frequency; NA – Not Applicable; R – Right; rTMS – repetitive Transcranial Magnetic Stimulation

### 3.4. Determinants of Antidepressant Effect

When considering continuous sociodemographic and clinical variables from the pooled RCT and uncontrolled data (Table S3), meta-regressions revealed that depression severity was significantly associated with greater antidepressant effect (β=0.24±0.08, p<0.005). On the other hand, longer illness duration was significantly associated to worse remission rates (β=-2.3±0.9, p<0.005; Table 3). We did not find significant associations between antidepressant effect and bipolar disease type, mean age, or female sex (Table 3). Meta-regression analysis also revealed that TMS was effective irrespective of inclusion of TRD patients in the studies (Table S8). Study quality (Table 3) and study design (Table S8) were not significant predictors of antidepressant effect.

**Table 3.** Meta-regression for continuous variables.

| Continuous Variables | Response rate |  |  | Remission rate |  |  | Dep. improvement |  |  |
| --- | --- | --- | --- | --- | --- | --- | --- | --- | --- |
| | $\beta \pm SE$ | p | N | $\beta \pm SE$ | p | N | $\beta \pm SE$ | p | N |
| **Demographic** |  |  |  |  |  |  |  |  |  |
| Mean age (y) | -0.75±0.57 | 0.20 | 40 | -0.18±0.48 | 0.71 | 34 | -0.43±0.02 | 0.06 | 34 |
| Sex (% female) | 0.04±0.33 | 0.91 | 36 | -0.32±0.37 | 0.41 | 31 | 0.01±0.13 | 0.41 | 33 |
| **Clinical** |  |  |  |  |  |  |  |  |  |
| BD-I (%) | -0.26±0.43 | 0.56 | 15 | -0.26±0.32 | 0.43 | 11 | -0.009±0.14 | 0.52 | 12 |
| Dep. duration (mo) | 1.02±1.49 | 0.51 | 12 | -0.12±1.5 | 0.94 | 10 | 0.02±0.06 | 0.71 | 13 |
| No. previous Dep. episodes | -5.07±2.56 | 0.10 | 8 | -0.32±2.77 | 0.91 | 7 | -0.27±0.14 | 0.10 | 8 |
| BD duration (y) | -0.81±1.17 | 0.50 | 17 | **-2.26±0.92** | **0.029** | **16** | -0.07±0.06 | 0.32 | 14 |
| Dep. Severity* | 3.54±2.71 | 0.201 | 37 | -2.11±2.28 | 0.36 | 33 | **0.24±0.08** | **0.004** | **39** |
| **Transcranial Magnetic Stimulation** |  |  |  |  |  |  |  |  |  |
| No. sessions | 0.90±0.52 | 0.09 | 52 | **1.07±0.51** | **0.046** | **34** | 0.05±0.03 | 0.07 | 33 |
| No. Trains | 0.13±0.11 | 0.25 | 32 | 0.18±0.09 | 0.07 | 22 | 0.003±0.005 | 0.48 | 20 |
| Inter-train Interval | -0.005±0.04 | 0.91 | 35 | -0.36±0.28 | 0.22 | 25 | -0.02±0.13 | 0.10 | 23 |
| Pulses/session | 0.005±0.004 | 0.20 | 49 | <0.001±<0.001 | 0.752 | 33 | <0.001±<0.001 | 0.93 | 31 |
| Stimulation Intensity | 0.14±0.30 | 0.64 | 48 | 0.29±0.27 | 0.284 | 33 | -0.01±0.01 | 0.30 | 31 |
| **Study Characteristics** |  |  |  |  |  |  |  |  |  |
| Study quality | -9.47±8.86 | 0.29 | 58 | -14.46±7.37 | 0.06 | 40 | 0.06±0.31 | 0.85 | 39 |

BD - bipolar disorder; BD-I - bipolar disorder type I; Dep. - Depression episode; Freq. - Frequency; H - Hemisphere; mo - months; n.s. - not significant; No. - Number; SE - Standard error; TRD - Treatment resistant depression; y – years
\* severity assessed with baseline depression rating scale normalized to standard deviation to make each study comparable among each other

When considering continuous TMS parameters, meta-regressions revealed that the number of TMS sessions was significantly associated with higher remission rates (β=1.1±0.5, p<0.005; Table 3). We did not find significant associations between antidepressant effect and number of pulses or trains per session, inter-train interval, and stimulation intensity (Table 3). Meta-regression analyses for binary TMS parameters showed that this is an effective antidepressant treatment irrespective of method to determine motor threshold, targeting method, use of accelerated protocols, frequency, laterality and side of stimulation (Table S8).

## 4. Discussion

Transcranial magnetic stimulation is a well-established treatment for several psychiatric disorders, based on consolidated regulatory approvals and evidenced-based guidelines(7). Nonetheless, and despite meta-analyses indicating that it is efficacious and safe, the use of TMS in BDep is still questioned(16, 17), largely because of conflicting results regarding efficacy in sham-controlled trials(13, 15). Despite originating in large part from small samples, this balance has fueled the debate regarding the place of rTMS in the management of BDep regardless of real-world data attesting its safety and efficacy(19, 64). Contributing towards the determination of the role of TMS for treatment of BDep, our data provides definitive evidence of a statistically significant effect over sham supporting efficacy, and of a large effect size (>0.8) supporting therapeutic effectiveness(77). This is, to our knowledge, the largest dataset to date yielding response and remission rates (47% and 27%, respectively), that are similar to those reported for MDD(78, 79). Importantly, we have also, for the first time, identified clear stimulation parameters and clinical characteristics associated with enhanced antidepressant effects in this population.

Our results confirm a clear and significant effect of TMS on depressive symptom severity in patients with BDep, similar to what has been found in unipolar depression(78). However, contrary to existing data establishing durability of treatment effects in MDD(80), we found insufficient data to appropriately determine how long after TMS antidepressant effects will last in BDep. This is particularly important given reports of transient effects in a large RCT, showing that active TMS was superior to sham at end point (4 weeks) but not at follow-up (8 weeks)(13). In contrast, Rapinesi *et al* (2018) provided 6-month follow-up data to support sustained antidepressant effects of rTMS in BDep, while Koutsumitros et al (2022) showed rates of 78% symptom remission at 1-month follow-up. In the future, studies evaluating the durability of antidepressant effect in BDep should be conducted to complement existing knowledge. We also extracted data concerning tolerability and safety. As in unipolar depression(81), the data collected here supports that TMS is well tolerated in BDep. The most common side effects were mild and transient, with a single uncomplicated generalized seizure reported in one paper, and treatment-emergent mania/hypomania was not significantly different between active and sham groups. These findings agree with existing evidence highlighting an excellent safety profile for rTMS, with extremely low rates of TMS-induced seizures or other severe side-effects, and mostly mild side effects being reported(81–83).

Although previous work had already suggested that rTMS is efficacious(16, 17, 84), our work extends beyond available systematic reviews and meta-analyses. In addition to confirming that rTMS is useful and safe for clinical management of BDep in the largest pool of studies and patients to date, we also explored, for the first time, potential predictors of antidepressant effect that may guide current clinical work and future research, similarly to what has been performed for other psychiatric conditions(85). First, regarding the characteristics of the treated population, studies including patients with more severe depressive symptoms at baseline were associated with better TMS antidepressant effects. While this finding is not surprising, since patients with the most severe symptoms have more room for measurable improvement(86), this has not been consistently reported in unipolar depression, for whom severity has been considered both a negative and a positive predictor of TMS effect(87). Regarding treatment resistance, while this is a negative predictor of response to rTMS in unipolar depression(88), we did not find equivalent evidence for BDep, where studies that included patients with TRD had similar antidepressant effect to those excluding this population. If confirmed, these differences reinforce important clinical distinctions between bipolar and unipolar depression (89), further suggesting that, in bipolar disorder, mood stabilization circuits are distinct from those affected in MDD(25, 90, 91). On the other hand, similarly to MDD(87), longer illness duration was associated with worse treatment outcomes, highlighting the importance of early accurate diagnosis and decisive treatment for all patients presenting with depressive symptoms(92). Further attention should be devoted in future research to the role of depression severity as well as illness chronicity as predictors of response to TMS in BDep.

Within the description of study-level predictors of antidepressant effectiveness, our work is of further value for practicing rTMS physicians, since it provides guidance on the most effective TMS protocols for BDep. Greater number of sessions was found to be significantly associated with higher remission rates, suggesting that longer treatments are associated to better outcomes, possibly as a result of cumulative neuroplastic effects over time(93). Again, this is not entirely surprising, but rather re-assuring, as it has been found for other populations treated with rTMS for depression, for example in geriatric samples(94, 95). Finally, and most importantly, our findings can help define, also for the first time, which TMS protocols are better suited to treat

BDep. Indeed, we found that both HF-rTMS-L and LF-rTMS-R were robustly and significantly associated with clinical antidepressant effect across the different outcomes, as in unipolar depression(7). Importantly, and contrary to negative results in previous studies(15), iTBS protocols targeting the left DLPFC also showed effectiveness in the treatment of BDep, in accordance with effecs in unipolar depression(79). Furthermore, antidepressant effectiveness for BDep did not depend on the number of pulses or trains per session, inter-train interval, stimulation intensity as well as method to determine motor threshold, target method, use of accelerated protocols, frequency, laterality and side of stimulation. Future work would benefit from determining, within restricted parameter space, the optimal rTMS parameters for BDep, including the comparison of high-frequency rTMS with iTBS.

The interpretation and application of our findings should consider the limitations of this work. First, twenty-seven studies reported mixed samples, consisting of patients diagnosed with bipolar or unipolar depression, and potential selection bias of the participants with BDep cannot thus be fully excluded. Second, additional information, such as demographics, psychiatric history and disease characteristics were not consistently reported in the analyzed studies. This limited the inclusion of such variables in the meta-regressions and sub-group meta-analysis, potentially restricting the ability to explore the impact of TMS in subdomains of symptoms in BDep, for example sleep(96, 97). According to best practices, we have only performed such analyses when sufficient studies were available to draw meaningful conclusions(21). Third, selected studies apply a variety of rTMS protocols, resulting in high heterogeneity between studies. Such heterogeneity of TMS protocols is likely to be related to the relative novelty of TMS as therapeutic approach for BDep treatment, and, as the field continues to mature, the most appropriate stimulation parameters are expected to be further clarified. Here we employed meta-regression and sub-group meta-analysis to contribute towards this clarification(21). Another source of heterogeneity results from different outcome measures being employed across studies. To mitigate this limitation, we used standardized effect-sizes as well as random-effect meta-analysis, minimizing the potential impact of these differences. Finally, the possibility of publication bias cannot be excluded, and examination of trial registrations reveals numerous potentially unpublished trials, including industry sponsored trials (for instance NCT01566591). However, it is noteworthy that Trim-Fill Duval-Tweedie Test suggests increased OR for response and remission in BDep when including potentially omitted studies, increasing confidence on our results regarding efficacy.

In conclusion, we show that rTMS is associated with significant antidepressant effect relative to sham, as well as a very large therapeutic effect. Most importantly, the rates of clinical response and remission parallel those reported in unipolar depression, and this treatment is also well tolerated in BDep and in MDD. Our results extend beyond previous literature since we have found potential determinants of such therapeutic effect. We show that illness severity and long treatment courses predicted greater antidepressant effects in BDep. Finally, we help guiding clinicians practicing TMS by showing that high and low frequency protocols on left and right DLPFC, respectively, are robustly associated to positive outcomes as in unipolar depression, with iTBS to L-DLPFC being a promising option that should be further studied.

## Data Availability

All data produced in the present work are contained in the manuscript

https://drive.google.com/file/d/1N8RIEwOwFQF9rPX9-w4fP9_jtp9eojpr/view?usp=drive_link

## REFERENCES

1. Merikangas KR, Jin R, He JP, Kessler RC, Lee S, Sampson NA, et al. Prevalence and correlates of bipolar spectrum disorder in the world mental health survey initiative. Arch Gen Psychiatry. 2011;68(3):241–51.

2. Blanco C, Compton WM, Saha TD, Goldstein BI, Ruan WJ, Huang B, et al. Epidemiology of DSM-5 bipolar I disorder: Results from the National Epidemiologic Survey on Alcohol and Related Conditions - III. J Psychiatr Res. 2017;84:310–7.

3. McDonald KC, Bulloch AG, Duffy A, Bresee L, Williams JV, Lavorato DH, et al. Prevalence of Bipolar I and II Disorder in Canada. Can J Psychiatry. 2015;60(3):151–6.

4. Forte A, Baldessarini RJ, Tondo L, Vázquez GH, Pompili M, Girardi P. Long-term morbidity in bipolar-I, bipolar-II, and unipolar major depressive disorders. J Affect Disord. 2015;178:71–8.

5. Keck PE, Jr., Kessler RC, Ross R. Clinical and economic effects of unrecognized or inadequately treated bipolar disorder. J Psychiatr Pract. 2008;14 Suppl 2:31–8.

6. Yatham LN, Kennedy SH, Parikh SV, Schaffer A, Bond DJ, Frey BN, et al. Canadian Network for Mood and Anxiety Treatments (CANMAT) and International Society for Bipolar Disorders (ISBD) 2018 guidelines for the management of patients with bipolar disorder. Bipolar Disord. 2018;20(2):97–170.

7. Cotovio G, Ventura F, Rodrigues da Silva D, Pereira P, Oliveira-Maia AJ. Regulatory Clearance and Approval of Therapeutic Protocols of Transcranial Magnetic Stimulation for Psychiatric Disorders. Brain Sci. 2023;13(7).

8. Valero-Cabré A, Amengual JL, Stengel C, Pascual-Leone A, Coubard OA. Transcranial magnetic stimulation in basic and clinical neuroscience: A comprehensive review of fundamental principles and novel insights. Neurosci Biobehav Rev. 2017;83:381–404.

9. Wischnewski M, Schutter DJ. Efficacy and Time Course of Theta Burst Stimulation in Healthy Humans. Brain Stimul. 2015;8(4):685–92.

10. Chen JJ, Zhao LB, Liu YY, Fan SH, Xie P. Comparative efficacy and acceptability of electroconvulsive therapy versus repetitive transcranial magnetic stimulation for major depression: A systematic review and multiple-treatments meta-analysis. Behav Brain Res. 2017;320:30–6.

11. Zhang M, Mo J, Zhang H, Tang Y, Guo K, OuYang X, et al. Efficacy and tolerability of repetitive transcranial magnetic stimulation for late-life depression: A systematic review and meta-analysis. Journal of Affective Disorders. 2023;323:219–31.

12. Camprodon JA. Therapeutic Neuromodulation for Bipolar Disorder—The Case for Biomarker-Driven Treatment Development. JAMA Network Open. 2021;4(3):e211055–e.

13. Tavares DF, Myczkowski ML, Alberto RL, Valiengo L, Rios RM, Gordon P, et al. Treatment of Bipolar Depression with Deep TMS: Results from a Double-Blind, Randomized, Parallel Group, Sham-Controlled Clinical Trial. Neuropsychopharmacology. 2017;42(13):2593–601.

14. Sheline YI, Makhoul W, Batzdorf AS, Nitchie FJ, Lynch KG, Cash R, et al. Accelerated Intermittent Theta-Burst Stimulation and Treatment-Refractory Bipolar Depression: A Randomized Clinical Trial. JAMA Psychiatry. 2024.

15. McGirr A, Vila-Rodriguez F, Cole J, Torres IJ, Arumugham SS, Keramatian K, et al. Efficacy of Active vs Sham Intermittent Theta Burst Transcranial Magnetic Stimulation for Patients With Bipolar Depression: A Randomized Clinical Trial. JAMA Netw Open. 2021;4(3):e210963.

16. Nguyen TD, Hieronymus F, Lorentzen R, McGirr A, Østergaard SD. The efficacy of repetitive transcranial magnetic stimulation (rTMS) for bipolar depression: A systematic review and meta-analysis. J Affect Disord. 2021;279:250–5.

17. Tee MMK, Au CH. A Systematic Review and Meta-Analysis of Randomized Sham-Controlled Trials of Repetitive Transcranial Magnetic Stimulation for Bipolar Disorder. Psychiatr Q. 2020;91(4):1225–47.

18. McGirr A, Karmani S, Arsappa R, Berlim MT, Thirthalli J, Muralidharan K, et al. Clinical efficacy and safety of repetitive transcranial magnetic stimulation in acute bipolar depression. World Psychiatry. 15. Italy2016. p. 85–6.

19. Gama-Chonlon L, Scanlan JM, Allen RM. Could bipolar depressed patients respond better to rTMS than unipolar depressed patients? A naturalistic, observational study. Psychiatry Research. 2022;312.

20. Yang YB, Chan P, Rayani K, McGirr A. Comparative Effectiveness of Repetitive Transcranial Magnetic Stimulation in Unipolar and Bipolar Depression. Can J Psychiatry. 66. United States2021. p. 313–5.

21. Higgins J, Thomas J, Chandler J, Cumpston M, Li T, Page M, et al. Cochrane Handbook for Systematic Reviews of Inter ventions version 6.4 (updated August 2023): Cochrane; 2023.

22. Liberati A, Altman DG, Tetzlaff J, Mulrow C, Gøtzsche PC, Ioannidis JP, et al. The PRISMA statement for reporting systematic reviews and meta-analyses of studies that evaluate health care interventions: explanation and elaboration. PLoS Med. 2009;6(7):e1000100.

23. Sterne JAC, Savović J, Page MJ, Elbers RG, Blencowe NS, Boutron I, et al. RoB 2: a revised tool for assessing risk of bias in randomised trials. Bmj. 2019;366:l4898.

24. Wells G, Shea B, O’Connell D, Peterson j, Welch V, Losos M, et al. The Newcastle– Ottawa Scale (NOS) for Assessing the Quality of Non-Randomized Studies in Meta-Analysis. . 2000; .

25. Cotovio G, Rodrigues da Silva D, Real Lage E, Seybert C, Oliveira-Maia AJ. Hemispheric asymmetry of motor cortex excitability in mood disorders - Evidence from a systematic review and meta-analysis. Clin Neurophysiol. 2022;137:25–37.

26. Doorsamy W, Joel Oluwaseye L. A Review of Missing Data Handling Techniques for Machine Learning. International Journal of Innovate Technology and Interdisciplinar Sciences; 2022. p. 971–1005.

27. Weir CJ, Butcher I, Assi V, Lewis SC, Murray GD, Langhorne P, et al. Dealing with missing standard deviation and mean values in meta-analysis of continuous outcomes: a systematic review. BMC Med Res Methodol. 2018;18(1):25.

28. Zabriskie BN, Cole N, Baldauf J, Decker C. The impact of correction methods on rare-event meta-analysis. Res Synth Methods. 2024;15(1):130–51.

29. Borenstein M. Comprehensive Meta-Analysis Software. 2022. p. 535–48.

30. Dolberg OT, Dannon PN, Schreiber S, Grunhaus L. Transcranial magnetic stimulation in patients with bipolar depression: a double blind, controlled study. Bipolar Disord. 2002;4 Suppl 1:94–5.

31. Nahas Z, Kozel FA, Li X, Anderson B, George MS. Left prefrontal transcranial magnetic stimulation (TMS) treatment of depression in bipolar affective disorder: a pilot study of acute safety and efficacy. Bipolar Disord. 2003;5(1):40–7.

32. Beynel L, Chauvin A, Guyader N, Harquel S, Szekely D, Bougerol T, et al. What saccadic eye movements tell us about TMS-induced neuromodulation of the DLPFC and mood changes: a pilot study in bipolar disorders. Front Integr Neurosci. 2014;8:65.

33. Fitzgerald PB, Hoy KE, Elliot D, McQueen S, Wambeek LE, Daskalakis ZJ. A negative double-blind controlled trial of sequential bilateral rTMS in the treatment of bipolar depression. J Affect Disord. 2016;198:158–62.

34. Hu SH, Lai JB, Xu DR, Qi HL, Peterson BS, Bao AM, et al. Efficacy of repetitive transcranial magnetic stimulation with quetiapine in treating bipolar II depression: a randomized, double-blinded, control study. Sci Rep. 2016;6:30537.

35. Bulteau S, Beynel L, Marendaz C, Dall’Igna G, Peré M, Harquel S, et al. Twice-daily neuronavigated intermittent theta burst stimulation for bipolar depression: A Randomized Sham-Controlled Pilot Study. Neurophysiol Clin. 2019;49(5):371–5.

36. Zengin G, Topak OZ, Atesci O, Culha Atesci F. The Efficacy and Safety of Transcranial Magnetic Stimulation in Treatment-Resistant Bipolar Depression. Psychiatr Danub. 2022;34(2):236–44.

37. Fitzgerald PB, Benitez J, de Castella A, Daskalakis ZJ, Brown TL, Kulkarni J. A randomized, controlled trial of sequential bilateral repetitive transcranial magnetic stimulation for treatment-resistant depression. Am J Psychiatry. 2006;163(1):88–94.

38. Herwig U, Fallgatter AJ, Höppner J, Eschweiler GW, Kron M, Hajak G, et al. Antidepressant effects of augmentative transcranial magnetic stimulation: randomised multicentre trial. Br J Psychiatry. 2007;191:441–8.

39. Speer AM, Wassermann EM, Benson BE, Herscovitch P, Post RM. Antidepressant efficacy of high and low frequency rTMS at 110% of motor threshold versus sham stimulation over left prefrontal cortex. Brain Stimul. 2014;7(1):36–41.

40. Kimbrell TA, Little JT, Dunn RT, Frye MA, Greenberg BD, Wassermann EM, et al. Frequency dependence of antidepressant response to left prefrontal repetitive transcranial magnetic stimulation (rTMS) as a function of baseline cerebral glucose metabolism. Biol Psychiatry. 1999;46(12):1603–13.

41. Klein E, Kreinin I, Chistyakov A, Koren D, Mecz L, Marmur S, et al. Therapeutic efficacy of right prefrontal slow repetitive transcranial magnetic stimulation in major depression: a double-blind controlled study. Arch Gen Psychiatry. 1999;56(4):315–20.

42. George MS, Nahas Z, Molloy M, Speer AM, Oliver NC, Li XB, et al. A controlled trial of daily left prefrontal cortex TMS for treating depression. Biol Psychiatry. 2000;48(10):962–70.

43. Loo CK, Mitchell PB, Croker VM, Malhi GS, Wen W, Gandevia SC, et al. Double-blind controlled investigation of bilateral prefrontal transcranial magnetic stimulation for the treatment of resistant major depression. Psychol Med. 2003;33(1):33–40.

44. Rossini D, Lucca A, Zanardi R, Magri L, Smeraldi E. Transcranial magnetic stimulation in treatment-resistant depressed patients: a double-blind, placebo-controlled trial. Psychiatry Res. 2005;137(1-2):1–10.

45. Su TP, Huang CC, Wei IH. Add-on rTMS for medication-resistant depression: a randomized, double-blind, sham-controlled trial in Chinese patients. J Clin Psychiatry. 2005;66(7):930–7.

46. McDonald WM, Easley K, Byrd EH, Holtzheimer P, Tuohy S, Woodard JL, et al. Combination rapid transcranial magnetic stimulation in treatment refractory depression. Neuropsychiatr Dis Treat. 2006;2(1):85–94.

47. Paillère Martinot ML, Galinowski A, Ringuenet D, Gallarda T, Lefaucheur JP, Bellivier F, et al. Influence of prefrontal target region on the efficacy of repetitive transcranial magnetic stimulation in patients with medication-resistant depression: a [(18)F]-fluorodeoxyglucose PET and MRI study. Int J Neuropsychopharmacol. 2010;13(1):45–59.

48. Hernández-Ribas R, Deus J, Pujol J, Segalàs C, Vallejo J, Menchón JM, et al. Identifying brain imaging correlates of clinical response to repetitive transcranial magnetic stimulation (rTMS) in major depression. Brain Stimul. 2013;6(1):54–61.

49. Chistyakov AV, Kreinin B, Marmor S, Kaplan B, Khatib A, Darawsheh N, et al. Preliminary assessment of the therapeutic efficacy of continuous theta-burst magnetic stimulation (cTBS) in major depression: a double-blind sham-controlled study. J Affect Disord. 2015;170:225–9.

50. Prasser J, Schecklmann M, Poeppl TB, Frank E, Kreuzer PM, Hajak G, et al. Bilateral prefrontal rTMS and theta burst TMS as an add-on treatment for depression: a randomized placebo controlled trial. World J Biol Psychiatry. 2015;16(1):57–65.

51. Mak ADP, Neggers SFW, Leung ONW, Chu WCW, Ho JYM, Chou IWY, et al. Antidepressant efficacy of low-frequency repetitive transcranial magnetic stimulation in antidepressant-nonresponding bipolar depression: a single-blind randomized sham-controlled trial. Int J Bipolar Disord. 2021;9(1):40.

52. Novák T, Kostýlková L, Bareš M, Renková V, Hejzlar M, Renka J, et al. Right ventrolateral and left dorsolateral 10 Hz transcranial magnetic stimulation as an add-on treatment for bipolar I and II depression: a double-blind, randomised, three-arm, sham-controlled study. World J Biol Psychiatry. 2024;25(5):304–16.

53. Mallik G, Mishra P, Garg S, Dhyani M, Tikka SK, Tyagi P. Safety and Efficacy of Continuous Theta Burst “Intensive” Stimulation in Acute-Phase Bipolar Depression A Pilot, Exploratory Study. Journal of Ect. 2023;39(1):28–33.

54. Dellink A, Hebbrecht K, Zeeuws D, Baeken C, De Fré G, Bervoets C, et al. Continuous theta burst stimulation for bipolar depression: A multicenter, double-blind randomized controlled study exploring treatment efficacy and predictive potential of kynurenine metabolites. J Affect Disord. 2024;361:693–701.

55. Ning L, Xue-Yi W, Zhen-Zhou Q, Mei S. A 4-week single-blind randomized controlled trial of repetitive transcranial magnetic stimulation combined with lithium and quetiapine in treatment of patients with bipolar depression. Chinese mental health journal. 2013;27(12):896–900.

56. Carnell BL, Clarke P, Gill S, Galletly CA. How effective is repetitive transcranial magnetic stimulation for bipolar depression? J Affect Disord. 2017;209:270–2.

57. Fitzgerald PB, Huntsman S, Gunewardene R, Kulkarni J, Daskalakis ZJ. A randomized trial of low-frequency right-prefrontal-cortex transcranial magnetic stimulation as augmentation in treatment-resistant major depression. Int J Neuropsychopharmacol. 2006;9(6):655–66.

58. Kito S, Miyazi M, Nakatani H, Matsuda Y, Yamazaki R, Okamoto T, et al. Effectiveness of high-frequency left prefrontal repetitive transcranial magnetic stimulation in patients with treatment-resistant depression: A randomized clinical trial of 37.5-minute vs 18.75-minute protocol. Neuropsychopharmacol Rep. 2019;39(3):203–8.

59. Olejarczyk E, Zuchowicz U, Wozniak-Kwasniewska A, Kaminski M, Szekely D, David O. The Impact of Repetitive Transcranial Magnetic Stimulation on Functional Connectivity in Major Depressive Disorder and Bipolar Disorder Evaluated by Directed Transfer Function and Indices Based on Graph Theory. Int J Neural Syst. 2020;30(4):2050015.

60. Rapinesi C, Kotzalidis GD, Ferracuti S, Girardi N, Zangen A, Sani G, et al. Add-on high frequency deep transcranial magnetic stimulation (dTMS) to bilateral prefrontal cortex in depressive episodes of patients with major depressive disorder, bipolar disorder I, and major depressive with alcohol use disorders. Neurosci Lett. 2018;671:128–32.

61. Rostami R, Kazemi R, Nitsche MA, Gholipour F, Salehinejad MA. Clinical and demographic predictors of response to rTMS treatment in unipolar and bipolar depressive disorders. Clin Neurophysiol. 2017;128(10):1961–70.

62. Harel EV, Zangen A, Roth Y, Reti I, Braw Y, Levkovitz Y. H-coil repetitive transcranial magnetic stimulation for the treatment of bipolar depression: an add-on, safety and feasibility study. World J Biol Psychiatry. 2011;12(2):119–26.

63. Dell’Osso B, Mundo E, D’Urso N, Pozzoli S, Buoli M, Ciabatti M, et al. Augmentative repetitive navigated transcranial magnetic stimulation (rTMS) in drug-resistant bipolar depression. Bipolar Disord. 2009;11(1):76–81.

64. Aaronson ST, Goldwaser EL, Croarkin PE, Geske JR, LeMahieu A, Sklar JH, et al. A Pilot Study of High-Frequency Transcranial Magnetic Stimulation for Bipolar Depression. J Clin Psychiatry. 2024;85(2).

65. Kazemi R, Rostami R, Khomami S, Baghdadi G, Rezaei M, Hata M, et al. Bilateral Transcranial Magnetic Stimulation on DLPFC Changes Resting State Networks and Cognitive Function in Patients With Bipolar Depression. Front Hum Neurosci. 2018;12:356.

66. Kazemi R, Rostami R, Khomami S, Horacek J, Brunovsky M, Novak T, et al. Electrophysiological correlates of bilateral and unilateral repetitive transcranial magnetic stimulation in patients with bipolar depression. Psychiatry Res. 2016;240:364–75.

67. Koutsomitros T, van der Zee KT, Evagorou O, Schuhmann T, Zamar AC, Sack AT. A Different rTMS Protocol for a Different Type of Depression: 20.000 rTMS Pulses for the Treatment of Bipolar Depression Type II. Journal of Clinical Medicine. 2022;11(18).

68. Alhelali A, Almheiri E, Abdelnaim M, Weber FC, Langguth B, Schecklmann M, et al. Effectiveness of Repetitive Transcranial Magnetic Stimulation in the Treatment of Bipolar Disorder in Comparison to the Treatment of Unipolar Depression in a Naturalistic Setting. Brain Sci. 2022;12(3).

69. Desbeaumes Jodoin V, Miron JP, Lespérance P. Safety and Efficacy of Accelerated Repetitive Transcranial Magnetic Stimulation Protocol in Elderly Depressed Unipolar and Bipolar Patients. Am J Geriatr Psychiatry. 2019;27(5):548–58.

70. Ikawa H, Osawa R, Takeda Y, Sato A, Mizuno H, Noda Y. Real-world retrospective study of repetitive transcranial magnetic stimulation (TMS) treatment for bipolar and unipolar depression using TMS registry data in Tokyo. Heliyon. 2024;10(5):e27288.

71. Bouaziz N, Laidi C, Bulteau S, Berjamin C, Thomas F, Moulier V, et al. Real world transcranial magnetic stimulation for major depression: A multisite, naturalistic, retrospective study. J Affect Disord. 2023;326:26–35.

72. Goldwaser EL, Daddario K, Aaronson ST. A retrospective analysis of bipolar depression treated with transcranial magnetic stimulation. Brain Behav. 2020;10(12):e01805.

73. Phillips AL, Burr RL, Dunner DL. Repetitive Transcranial Magnetic Stimulation in the Treatment of Bipolar Depression: Experience From a Clinical Setting. J Psychiatr Pract. 2020;26(1):37–45.

74. Fitzgerald PB, Brown TL, Marston NA, Daskalakis ZJ, De Castella A, Kulkarni J. Transcranial magnetic stimulation in the treatment of depression: a double-blind, placebo-controlled trial. Arch Gen Psychiatry. 2003;60(10):1002–8.

75. Dalton JE, Bolen SD, Mascha EJ. Publication Bias: The Elephant in the Review. Anesth Analg. 123. United States2016. p. 812–3.

76. Bretlau LG, Lunde M, Lindberg L, Undén M, Dissing S, Bech P. Repetitive transcranial magnetic stimulation (rTMS) in combination with escitalopram in patients with treatment-resistant major depression: a double-blind, randomised, sham-controlled trial. Pharmacopsychiatry. 2008;41(2):41–7.

77. Serdar CC, Cihan M, Yücel D, Serdar MA. Sample size, power and effect size revisited: simplified and practical approaches in pre-clinical, clinical and laboratory studies. Biochem Med (Zagreb). 2021;31(1):010502.

78. Thompson L. Treating major depression and comorbid disorders with transcranial magnetic stimulation. J Affect Disord. 2020;276:453–60.

79. Rush AJ, Trivedi MH, Wisniewski SR, Nierenberg AA, Stewart JW, Warden D, et al. Acute and longer-term outcomes in depressed outpatients requiring one or several treatment steps: a STAR*D report. Am J Psychiatry. 2006;163(11):1905–17.

80. Senova S, Cotovio G, Pascual-Leone A, Oliveira-Maia AJ. Durability of antidepressant response to repetitive transcranial magnetic stimulation: Systematic review and meta-analysis. Brain Stimul. 2019;12(1):119–28.

81. Rossi S, Antal A, Bestmann S, Bikson M, Brewer C, Brockmöller J, et al. Safety and recommendations for TMS use in healthy subjects and patient populations, with updates on training, ethical and regulatory issues: Expert Guidelines. Clin Neurophysiol. 2021;132(1):269–306.

82. Stultz DJ, Osburn S, Burns T, Pawlowska-Wajswol S, Walton R. Transcranial Magnetic Stimulation (TMS) Safety with Respect to Seizures: A Literature Review. Neuropsychiatr Dis Treat. 2020;16:2989–3000.

83. Miuli A, Sepede G, Stigliano G, Mosca A, Di Carlo F, d’Andrea G, et al. Hypomanic/manic switch after transcranial magnetic stimulation in mood disorders: A systematic review and meta-analysis. World J Psychiatry. 2021;11(8):477–90.

84. Kishi T, Ikuta T, Sakuma K, Hatano M, Matsuda Y, Kito S, et al. Repetitive transcranial magnetic stimulation for bipolar depression: a systematic review and pairwise and network meta-analysis. Mol Psychiatry. 2024;29(1):39–42.

85. Steuber ER, McGuire JF. A Meta-analysis of Transcranial Magnetic Stimulation in Obsessive-Compulsive Disorder. Biol Psychiatry Cogn Neurosci Neuroimaging. 2023;8(11):1145–55.

86. McAllister-Williams RH. Do antidepressants work? A commentary on “Initial severity and antidepressant benefits: a meta-analysis of data submitted to the Food and Drug Administration” by Kirsch et al. Evid Based Ment Health. 2008;11(3):66–8.

87. Kar SK. Predictors of Response to Repetitive Transcranial Magnetic Stimulation in Depression: A Review of Recent Updates. Clin Psychopharmacol Neurosci. 2019;17(1):25–33.

88. Li CT, Su TP, Cheng CM, Chen MH, Bai YM, Tsai SJ. Factors associated with antidepressant responses to repetitive transcranial magnetic stimulation in antidepressant-resistant depression. Front Neurosci. 2022;16:1046920.

89. Hirschfeld RM. Differential diagnosis of bipolar disorder and major depressive disorder. J Affect Disord. 2014;169 Suppl 1:S12–6.

90. Han KM, De Berardis D, Fornaro M, Kim YK. Differentiating between bipolar and unipolar depression in functional and structural MRI studies. Prog Neuropsychopharmacol Biol Psychiatry. 2019;91:20–7.

91. Cotovio G, Talmasov D, Barahona-Corrêa JB, Hsu J, Senova S, Ribeiro R, et al. Mapping mania symptoms based on focal brain damage. J Clin Invest. 2020;130(10):5209–22.

92. Poleszczyk A, Rakowicz M, Parnowski T, Antczak J, Święcicki Ł. Are there clinical and neurophysiologic predictive factors for a positive response to HF-rTMS in patients with treatment-resistant depression? Psychiatry Res. 2018;264:175–81.

93. Briley PM, Webster L, Lankappa S, Pszczolkowski S, McAllister-Williams RH, Liddle PF, et al. Trajectories of improvement with repetitive transcranial magnetic stimulation for treatment-resistant major depression in the BRIGhTMIND trial. npj Mental Health Research. 2024;3(1):32.

94. Valiengo L, Maia A, Cotovio G, Gordon PC, Brunoni AR, Forlenza OV, et al. Repetitive Transcranial Magnetic Stimulation for Major Depressive Disorder in Older Adults: Systematic Review and Meta-analysis. J Gerontol A Biol Sci Med Sci. 2022;77(4):851–60.

95. Cotovio G, Boes AD, Press DZ, Oliveira-Maia AJ, Pascual-Leone A. In Older Adults the Antidepressant Effect of Repetitive Transcranial Magnetic Stimulation Is Similar but Occurs Later Than in Younger Adults. Front Aging Neurosci. 2022;14:919734.

96. Guerrera CS, Boccaccio FM, Varrasi S, Platania GA, Coco M, Pirrone C, et al. A narrative review on insomnia and hypersomnolence within Major Depressive Disorder and bipolar disorder: A proposal for a novel psychometric protocol. Neurosci Biobehav Rev. 2024;158:105575.

97. Lanza G, Fisicaro F, Cantone M, Pennisi M, Cosentino FII, Lanuzza B, et al. Repetitive transcranial magnetic stimulation in primary sleep disorders. Sleep Med Rev. 2023;67:101735.

