## Supplementary material for "Efficacy, effectiveness and safety of transcranial magnetic stimulation for bipolar depression: A systematic review and meta-analysis"

#### Supplementary Tables

**Table S1 –** Search syntax.

| | Psychiatric Disorder | Non-invasive Brain Stimulation Modality | Not Topic |
|---|---|---|---|
| Pubmed* | depressive disorder | transcranial stimulation | |
| | resistant depression | TMS | |
| | depressive episode | transcranial magnetic stimulation | |
| | depression | Non-invasive brain stimulation | |
| | bipolar | NIBS | |
| | bipolar disorder | intermittent theta-burst stimulation | |
| | manic depressive | iTBS | |
| | | Theta-burst stimulation | |
| | | TBS | |
| | | continuous theta-burst stimulation | |
| | | cTBS | |

| | | | |
|---|---|---|---|
| | | Deep TMS<br>dTMS<br>repetitive transcranial magnetic stimulation<br>rTMS | |
| Web of Science** | depressive disorder<br>resistant depression<br>depressive episode<br>depression<br>bipolar<br>bipolar disorder<br>manic depressive | transcranial stimulation<br>TMS<br>transcranial magnetic stimulation<br>Non-invasive brain stimulation<br>NIBS<br>intermittent theta-burst stimulation<br>iTBS<br>Theta-burst stimulation<br>TBS<br>continuous theta-burst stimulation<br>cTBS | Animal<br>Monkey<br>Chimpanzee<br>Mouse<br>Mice<br>Rat<br>Cat<br>Dog<br>Rabbit<br>Bird<br>Fish |

| | | | |
|---|---|---|---|
| | | Deep TMS<br>dTMS<br>repetitive transcranial magnetic stimulation<br>rTMS | Child |
| EMBASE*** | depressive disorder<br>resistant depression<br>depressive episode<br>depression<br>bipolar<br>bipolar disorder<br>manic depressive | transcranial stimulation<br>TMS<br>transcranial magnetic stimulation<br>Non-invasive brain stimulation<br>NIBS<br>intermittent theta-burst stimulation<br>iTBS<br>Theta-burst stimulation<br>TBS<br>continuous theta-burst stimulation<br>cTBS | |

| | | |
|---|---|---|
| | | Deep TMS |
| | | dTMS |
| | | repetitive transcranial magnetic stimulation |
| | | rTMS |
| Cochrane Library | depressive disorder | transcranial stimulation |
| | resistant depression | TMS |
| | depressive episode | transcranial magnetic stimulation |
| | depression | Non-invasive brain stimulation |
| | bipolar | NIBS |
| | bipolar disorder | intermittent theta-burst stimulation |
| | manic depressive | iTBS |
| | | Theta-burst stimulation |
| | | TBS |
| | | continuous theta-burst stimulation |
| | | cTBS |

| | |
|---|---|
| | Deep TMS |
| | dTMS |
| | repetitive transcranial magnetic stimulation |
| | rTMS |

* We applied the following filters: Humans; age +19; English, Portuguese, Spanish, French and Chinese.

** We applied the following filters: English, Portuguese, Spanish, French and Chinese; Articles, Review articles.

*** We applied the following filters: Humans; Young adults, adult, middle aged, aged and very elderly.

**Table S2** – Data not reported in the available articles and sent by the authors.

| Author | Data sent |
|---|---|
| Alexander McGirr | Additional data on response in McGirr et al. 2021 (1)<br>Percentage of remission in Yang et al. 2021(2)<br>Number of responders in active and sham groups for bipolar patients in Kimbrell et al. 1999(3), Klein et al. 1999(4), George et al. 2000(5), Loo et al.2003(6), Rossini et al.2005(7), Su et al. 2005(8), McDonald et al. 2006(9), Pallière Martinot et al. 2010(10), Hernández-Ribas et al. 2013(11), Chistyakov et al. 2015(12), Prasser et al. 2015(13) |
| Bulteau et al. 2019(14) | Demographics and MARDS score on bipolar population |
| Herwig et al. 2007(15) | Demographics and BDI/HAM/MADRS score on bipolar population |
| Debaunes Jodoin et al. 2018(16) | Response and remission rates, MADRS score on bipolar population |
| Kito et al. 2019(17) | Demographics, response and remission rates on bipolar population |
| Bouaziz et al. 2023(18) | Response and remission rates, baseline and post-treatment MADRS for bipolar population |
| Dellink et al. 2024(19) | Remission rates for bipolar population |

**Table S3** – Clinical and demographic data of the studies.

| | | Total sample | BDI | BDII | Female | Age (years) | Length current episode (months) | No. depressive episodes | Duration illness (years) |
|---|---|---|---|---|---|---|---|---|---|
| | | N | N | N | % | Mean | Mean | Mean | Mean |
| Study | Sham-Control Study | Active \| Sham | Active \| Sham | Active \| Sham | Active \| Sham | Active \| Sham | Active \| Sham | Active \| Sham | Active \| Sham |
| Kimbrell et al. 1999 (20Hz) | Yes | 3 \| 1 | 2 \| 0 | 0 \| 1 | 0 \| 0 | 51 \| 28 | | | |
| Klein et al. 1999 | Yes | 7 \| 6 | | | | | | | |
| George at al. 2000 | Yes | 7 \| 2 | | | | | | | |
| Dolberg et al. 2002 | Yes | 10 \| 10 | | | 50.00 \| 30.00 | 51.80 \| 59.00 | 3.00 \| 4.80 | | |
| Loo et al. 2003 | Yes | 2 \| 1 | | | | | | | |
| Nahas et al. 2003 | Yes | 11 \| 12 | 14 | 9 | 63.63 \| 58.33 | 42.40 \| 43.40 | 18.60 \| 23.50 | | 22.60 \| 19.30 |
| Rossini et al. 2005 | Yes | 12 \| 5 | | | | | | | |
| Su et al. 2005 | Yes | 3 \| 2 | | | | | | | |
| Fitzgerald et al. 2006a | Yes | 4 \| 4 | 8 | 0 | | | | | |
| Fitzgerald et al. 2006b (1Hz) | No | 12 \| | 6 | 6 | | | | | |
| Fitzgerald et al. 2006b (2Hz) | No | 13 \| | 8 | 5 | | | | | |

| | | | | | | | | | |
|---|---|---|---|---|---|---|---|---|---|
| McDonald et al. 2006 | Yes | 5 \| 3 | | | | | | | |
| Herwig et al. 2007 | Yes | 7 \| 4 | | | | 48.00 \| 44.00 | | | 16.15 \| 20.30 |
| Dell Osso et al. 2009 | No | 11 \| | 5 | 6 | 72.70 \| | 54.36 \| | | | |
| Pallière Martinot et al. 2010 | Yes | 11 \| 5 | | | 54.55\| 60.00 | | | | |
| Harel et al. 2011 | No | 19 \| | | | 57.89 \| | 45.47 \| | 11.28 \| | 9.94 \| | 18.18 \| |
| Hernández-Ribas el al. 2013 | Yes | 5 \| 1 | | | | | | | |
| Ning et al. 2013 | Yes | 30 \| 29 | 14 \| 15 | 16 \| 14 | 40.00 \| 48.28 | 34.00 \| 33.00 | 4.40 \| 5.20 | 2.00 \| 2.00 | 11.00 \| 11.00 |
| Beynel et al. 2014 | Yes | 5 \| 7 | | | | | | | |
| Speer et al. 2014 (L-HF) | Yes | 6 \| 2 | 3 \| 1 | 3 \| 1 | 50.00 \| 0 | 38.30 \| 49.50 | | | |
| Chistyakov et al. 2015 | Yes | 6 \| 4 | | | | | | | |
| Prasser et al. 2015 (LF/HF) | Yes | 4 \| 6 | | | | | | | |
| Prasser et al. 2015 (cTBS/iTBS) | Yes | 8 \| 6 | | | | | | | |
| Carnell et al. 2016 | No | 50 \| | | | 66.00 \| | 48.40 \| | | | 23.00 \| |
| Fitzgerald et al. 2016 | Yes | 22 \| 23 | 8 \| 11 | 14 \| 12 | | 46.30 \| 49.70 | 3.50 \| 8.10 | | 20.10 \| 24.00 |
| Hu et al. 2016 (L-HF) | Yes | 11 \| 12 | | 11 \| 12 | 54.54 \| 58.33 | 27.40 \| 23.70 | | 1.64 \| 3.17 | 23.70 \| 20.30 |
| Hu et al. 2016 (R-LF) | Yes | 12 \| 12 | | 12 \| 12 | 41.60 \| 58.33 | 28.30 \| 23.70 | | 1.92 \| 3.17 | 24.50 \| 20.30 |

| | | | | | | | | |
|---|---|---|---|---|---|---|---|---|
| Kazemi et al. 2016 (R-LF & L-HF) | No | 15 \| | | | 53.00 \| | 34.67 \| | | |
| Kazemi et al. 2016 (R-LF) | No | 15 \| | | | 60.00 \| | 36.13 \| | | |
| Rostami et al. 2017 | No | 146 \| | | | 62.33 \| | 34.28 \| | | |
| Tavares et al. 2017 | Yes | 25 \| 25 | 11 \| 14 | 14 \| 11 | 68.00 \| 72.00 | 43.50 \| 41.20 | 6.00 \| 8.00 | |
| Debaunes Jodoin et al. 2018 | No | 16 \| | | | 56.25 \| | 44.87 \| | | |
| Kazemi et al. 2018 | No | 20 \| | | | 60.00 \| | 28.65 \| | | |
| Rapinesi et al. 2018 | No | 20 \| | 20 | | 45.00 \| | 57.85 \| | | |
| Bulteau et al. 2019 | Yes | 12 \| 14 | | | 58.33 \| 28.57 | 52.75 \| 53.14 | 17.30 \| 13.25 | |
| Kito et al. 2019 (ITI=11s) | No | 6 \| | | | | 43.30 \| | | 7.00 \| |
| Kito et al. 2019 (ITI=26s) | No | 5 \| | | | | 52.40 \| | | 15.60 \| |
| Goldwaser et al. 2020 | No | 39 \| | 18 | 21 | | | | |
| Olejarczyk et al. 2020 | No | 10 \| | | | | 48.70 \| | 18.48 \| | |
| Phillips et al. 2020 | No | 17 \| | 3 | 12 | 52.94 \| | 46.70 \| | | 24.80 \| |
| Mak et al. 2021 | Yes | 23 \| 25 | 4 \| 4 | 19 \| 21 | 65.22 \| 64.00 | 39.70 \| 40.0 | | 17.00 \| 16.30 |
| McGirr et al. 2021 | Yes | 18 \| 19 | 11 \| 10 | 7 \| 9 | 61.10 \| 63.20 | 44.78 \| 43.00 | 7.59 \| 7.33 | |
| Yang et al. 2021 | No | 13 \| | 7 | 6 | 38.46 \| | 54.00 \| | 5.30 \| | |

| | | | | | | | | | |
|---|---|---|---|---|---|---|---|---|---|
| Abdullah Alhelali et al. 2022 | No | 46 \| | | | 54.35 \| | 48.00 \| | | | |
| Gama-Chonlon et al. 2022 (10 Hz) | No | 20 \| | | | | | | | |
| Gama-Chonlon et al. 2022 (R-LF & L-HF) | No | 7 \| | | | | | | | |
| Gama-Chonlon et al. 2022 (mixed) | No | 7 \| | | | | | | | |
| Koutsomitros et al. 2022 | No | 23 \| | 0 \| | 23 \| | 52.20 \| | | | | |
| Zengin et al. 2022 | Yes | 14 \| 15 | 23 | 6 | 57.14 \| 46.70 | 42.36 \| 38.93 | | | 17.36 \| 13.53 |
| Bouaziz et al. 2023 (1Hz) | No | 3 \| | | | 100.00 \| | 56.00 \| | | | |
| Bouaziz et al. 2023 (10Hz) | No | 38 \| | | | 55.30 \| | 59.00 \| | | | |
| Bouaziz et al. 2023 (20Hz) | No | 20 \| | | | 50.00 \| | 51.50 \| | | | |
| Bouaziz et al. 2023 (iTBS) | No | 52 \| | | | 69.20 \| | 44.52 \| | | | |
| Aaronson et al. 2024 | No | 31 \| | | | 58.1 \| | 44.52 \| | | | |
| Mallik et al. 2023 | Yes | 11 \| 8 | | | 54.50 \| 37.50 | 43.00 \| 34.63 | 3.64 \| 3.50 | 3.27 \| 3.43 | |
| Ikawa et al. 2024 | No | 20 \| | | | 45.00 \| | 36.80 \| | | | 4.40 \| |
| Dellink et al. 2024 | Yes | 18 \| 19 | | | 72 \| 68 | 48 \| 51 | | | |
| Novak et al. 2024 (RVL) | Yes | 20 \| 20 | 14 \| 13 | 6 \|7 | 65 \| 60 | 39.3 \| 43.9 | 5.05 \| 6.72 | 5.6 \| 8.5 | 11.5\| 14.8 |

| | | | | | | | | | |
|---|---|---|---|---|---|---|---|---|---|
| Novak et al. 2024 (LDL) | Yes | 20 \| 20 | 17 \| 13 | 3 \| 7 | 60 \| 60 | 48.9 \| 43.9 | 4.65 \| 6.72 | 9.0 \| 8.5 | 17.5 \| 14.8 |
| Sheline t al. 2024 | Yes | 12 \| 12 | 1 \| 11 | 1 \| 11 | 50 \| 50 | 41.3 \| 43.6 | | | |

BDI - Bipolar Disorder type I; BDII - Bipolar Disorder type II; HF – High frequency; ITI - Inter-train interval; L – Left; LF – Low frequency; LDL – left dorsolateral; No. – Number; R – Right; RVL - right ventrolateral.

**Table S4** – Treatment resistant depression criteria and medication status for each study

| Authors, year | TRD criteria | Concurrent medications |
|---|---|---|
| Kimbrell et al. 1999 | | Patients were allowed to remain on mood stabilizer treatment (1 bipolar II patien (on lithium); and 3 bipolar I patients (one on lithium and carbamazepine, one on lithium and lamotrigine, and one medication free)). |
| Klein et al. 1999 | | |
| George at al. 2000 | | Patients were free of antidepressant medications for at least 2 weeks before study entry, although three bipolar patients required ongoing mood stabilizers or benzodiazapines for anxiety. |
| Dolberg et al. 2002 | | |
| Loo et al. 2003 | Failure to respond to one adequate course of antidepressants. | |
| Nahas et al. 2003 | | Subjects could only be on Carbamazepine or Valproate on a stable dose for the last 2 weeks. All other medication was tapered over a 2 week washout. |
| Rossini et al. 2005 | Lack of improvement to at least two different treatments with antidepressants. | All drugs were maintained at a stable dosage during the duration of the trial. Lithium carbonate were allowd in bipolar patients. |
| Su et al. 2005 | Patients who had failed to respond to at least 2 adequate trials of antidepressant medications prior to rTMS treatmen. | All patients continued their current antidepressant medications during the 2-week course of rTMS administration. No medication changes were allowed for at least 4 weeks preceding rTMS and throughout the period of rTMS treatment. |
| Fitzgerald et al. 2006a | No response to 2 cycles of antidepressant for 6 weeks. | Patients were not withdrawn from medication before the trial. Medications were not allowed to have changed in the 4 weeks prior to commencement of the trial or during the trial itself. |

| | | |
|---|---|---|
| Fitzgerald et al. 2006b | Failure to respond to a minimum 2 courses of medication for at least 6 weeks in the current episode. | Medications were not allowed to have changed in the 4 weeks prior to commencement of the trial or during the trial itself. |
| McDonald et al. 2006 | Treatment resistance to at least 3 antidepressant medications during the presente depressive episode (no significant improvement in depressive symptoms following a 6-week trial of an antidepressant dosage equivalent to fluoxetine 20 mg). | Subjects that required continued treatment with antidepressant medications were excluded. |
| Herwig et al. 2007 | Lack of response to 2 different antidepressants and 1 combination of 4 weeks in sufficient dosage in the current episode. | Prior antidepressant medication was washed out (4 t½). Venlafaxine was started at a dosage of 75mg per day in the first week, and mirtazapine at a dosage of 15mg per day. No other antidepressant or concomitant antipsychotic medication was allowed. |
| Dell Osso et al. 2009 | | Patients maintained on treatment, as long as it was stabilized for 6 weeks |
| Pallière Martinot et al. 2010 | Resistance to at least two trials of antidepressants of different classes given at adequate doses (>150 mg/d in an equivalent dose of imipramine) and duration (at least 4 wk for each drug). | During the study, the patients were treated with minimal and stable doses of their previous treatment for at least 2 wk. Low-dose hypnotics prescribed in a naturalistic manner were allowed in case of severe insomnia only. |
| Harel et al. 2011 | | Patients were taking antidepressants and mood stabilizers. |
| Hernández-Ribas el al. 2013 | No response to at least one trial of an adequate depression treatment. | Stable pharmacological treatment for at least 6 weeks, which remained unchanged during the study period. |
| Ning et al. 2013 | | Both groups were taking lithium and quetiapine. No antidepressants or other antipsychotic drugs were used. Clonazepam was given for insomnia and anxiety. |
| Beynel et al. 2014 | No response to any antidepressant for at least 4 weeks. | Mood stabilizers during treatment period only if patient was stable at least 4 weeks |

| | | |
|---|---|---|
| | | before rTMS treatment. |
| Speer et al. 2014 | Failure to respond to at least 2 previous antidepressant trials. | |
| Chistyakov et al. 2015 | | |
| Prasser et al. 2015 | | |
| Carnell et al. 2016 | | It was requested that medications not be changed during the course of rTMS. |
| Fitzgerald et al. 2016 | Failure to respond to a minimum 2 courses of medication for at least 6 weeks in the current episode. | No increase or initiation of new antidepressant medication 4 weeks prior to beginning of trial. No change of medication allowed during trial. |
| Hu et al. 2016 | | All patients are drug naive or withdrawn from medication for at least 1 week prior to commencement of the study. All patients received Quetiapine. |
| Kazemi et al. 2016 | | Benzodiazepines were not used by patients of both groups. All patients were on stable medication during the study (mood stabilizers, SSRI, TCA and atypical antipsychotics). |
| Rostami et al. 2017 | | Stable regimen of medications for 4 weeks before treatment, with no changes throughout the course of treatment. Benzodiazepines were not used by any patient of both treatment groups. |
| Tavares et al. 2017 | Failure to achieve remission with ⩾2 interventions approved as first, second or third line therapies according to CANMAT | No antidepressant medication. |

| | | |
|---|---|---|
| | guidelines. | |
| Debaunes Jodoin et al. 2018 | Two trials of antidepressant medication of adequate dosing and duration. | |
| Kazemi et al. 2018 | | Unchanged medication regime during the treatment process. |
| Rapinesi et al. 2018 | Failure to respond to current medication for at least 1 month. | Treatment remained unchanged for the entire duration of the study. |
| Bulteau et al. 2019 | Failure of mood-stabilizing treatment at adequate duration and dosage for at least 1 month. | Stable mono or bi-therapy with mood stabilizers and without antidepressants nor benzodiazepines. |
| Kito et al. 2019 | | Medications given to the patients were not allowed to have changed in the 4 weeks before the start of the first rTMS treatment or during the trial. |
| Goldwaser et al. 2020 | | At least one mood stabilizer at an effective dose for at least two weeks and were taken off antidepressants prior to TMS treatment. |
| Olejarczyk et al. 2020 | No response to pharmacological therapy using minimum two distinctly different classes of antidepressant medications for actual depressive episode. | Mood stabilizer medication has been optimized according to recent recommendation and remained the same for at least two weeks prior to the entry in the study and during the study. Only cyanemazine and hydroxyzine, and no benzodiazepines, were used during this period. |
| Phillips et al. 2020 | Failure to respond to at least 2 antidepressants or 2 adjunctive therapies or had experienced intolerable side effects from antidepressant medications. | |

| | | |
|---|---|---|
| Mak et al. 2021 | No response to at least one previous adequate antidepressant (CANMAT 2013 guidelines) for at least 6 weeks in addition to at least one mood stabilizer. | Alterations to medication during trial were avoided. |
| McGirr et al. 2021 | No clinical response to at least 1 CANMAT-recommended first-line treatment for an acute episode. | Patients who had been taking antidepressant medications were allowed to continue these at same dose levels during the double-blind phase of the study. |
| Yang et al. 2021 | | Patients remained on stable pharmacological regimens throughout their treatments. |
| Abdullah Alhelali et al. 2022 | | |
| Gama-Chonlon et al. 2022 | | The majority of bipolar patients were prescribed mood stabilizers (26). 21 patients were on antidepressants, 20 were on antipsychotics, 13 were on lithium and 13 were on benzodiazepines. Medications were adjusted during the treatment course in some cases. |
| Koutsomitros et al. 2022 | | Patients were all on a mood stabilizer, mostly second-generation antipsychotics. Most of the patients were either on Quetiapine (9 patients) or Olanzapine (8 patients) monotherapy without any other mood stabilizer. Five patients were on Lithium, and six patients were also using an antidepressant: five were on SSRIs (Fluoxetine or Escitalopram), and one was on SNRI (Venlafaxine). None of the eleven patients on benzodiazepines used more than 4 mg equivalent of lorazepam per day. |
| Zengin et al. 2022 | No response to ≥2 interventions approved as first, second or | Mood-regulating drugs serum levels in the treatment range, and that no change during |

| | | |
|---|---|---|
| | third stage treatments according to CANMAT. | the trial. |
| Bouaziz et al. 2023 | Resistance to, or intolerance of, at least two antidepressant trials of adequate doses and duration. | |
| Mallik et al. 2023 | | Continue the psychotropic medication at the same dosages for the entire duration of the trial. |
| Aaronson et al. 2024 | | All patients were required to be on a suitable mood stabilizer (or/including) antipsychotic medication clinically used for bipolar disorder, as determined by the study clinician, for at least 4 days prior to the commencement of rTMS treatment. |
| Ikawa et al. 2024 | Maudsley staging method = 6.7 (±1.1) | |
| Dellink et al. 2024 | | Stable psychotropic regimen at least 2 weeks prior to randomization. During the study period, no changes to patients' psychotropic medication regimen were allowed. |
| Novak et al. 2024 | Failed to respond to at least one adequate treatment trial with mood stabilizers or add-on antidepressants in the current BDE. | A stable and adequate dose of mood stabilizers or antipsychotics for at least 4 weeks was required prior to screening. Lithium: RVL=8 (40%), LDL=8 (40%), S=5 (25%); Valproate: RVL=7 (35%), LDL=8 (40%), S=7 (35%); Lamotrigine: RVL=3 (15%), LDL=2 (10%), S=7 (35%); Antipsychotics: RVL=11 (55%), LDL=15 (75%), S=12 (60%); Antidepressants: RVL=15 (75%), LDL=13 (65%), S=15 (75%). The concomitant medications allowed in cases of insomnia or anxiety were z-hypnotics (zolpidem up to 10 mg, or zopiclone 7.5 mg at night), hydroxyzine (25 mg, 50 mg dose per day), and clonazepam (0.5 mg per dose, 1 mg per day). |

| | | |
|---|---|---|
| Sheline t al. 2024 | 2 or more failed treatments by Antidepressant Treatment History Form criteria. | Stable mood stabilizer regimen 4 or more weeks prior to aiTBS. |

CANMAT - Canadian Network for Mood and Anxiety Treatments; rTMS – repetitive Transcranial Magnetic Stimulation; TMS - Transcranial Magnetic Stimulation; TRD - Treatment-resistant depression.

**Table S5** - Repetitive Transcranial Magnetic Stimulation (rTMS) characteristics of the studies.

| Study | Stimulator brand | Coil type | Frequency (Hz) | Intensity | | Stimulation | | | No. of sessions | No. of sessions per day | No. of pulses per session | Trains (N) | ITI (sec) | Stim protocol |
|---|---|---|---|---|---|---|---|---|---|---|---|---|---|---|
| | | | | % rMT | Method | Target | Side | Method | | | | | | |
| Kimbrell et al. 1999 (20Hz) | Cadwell High Speed | figure-eight coil | 20 | 80 | EMG | DLPFC | Left | 5 cm | 10 | 1 | 800 | 20 | 60 | HF-rTMS-L |
| Klein et al. 1999 | Caldwell | circular coil | 1 | 110 | | DLPFC | Right | 6 cm | 10 | 1 | 120 | 2 | 180 | LF-rTMS-R |
| George et al. 2000 | Caldwell | figure-eight coil | 5/20 | 100 | | DLPFC | left | 5 cm | 10 | 1 | 1600 | | | |
| Dolberg et al. 2002 | | | | | | | | | 20 | | | | | |
| Loo et al. 2003 | MagStim Rapid | figure-eight coil | 15 | 90 | EMG | DLPFC | BL | 5 cm | 15 | 1 | 1800 | 24 | 25 | HF-rTMS-BL |
| Nahas et al. 2003 | Neotonus, Inc | figure-eight coil | 5 | 110 | visual | DLPFC | Left | 5 cm | 10 | 1 | 1600 | 40 | 22 | HF-rTMS-L |
| Rossini et al. 2005 | MagStim Rapid | figure-eight coil | 15 | 80/100 | visual | DLPFC | Left | 5 cm | 10 | 1 | 600 | 20 | 28 | HF-rTMS-L |
| Su et al. 2005 | MagStim Rapid | figure-eight coil | 5/20 | 100 | | DLPFC | Left | 5 cm | 10 | 1 | 1600 | 40 | | |
| Fitzgerald et al. 2006a | MagProX100, MagVenture | figure-eight coil | R -1<br>L - 10 | R- 110<br>L -100 | EMG | DLPFC | BL | 5 cm | 10 | 1 | | R -3<br>L- 15 | R- 30<br>L- 25 | LF-rTMS-R followed by HF-rTMS-L |

| | | | | | | | | | | | | | | |
|---|---|---|---|---|---|---|---|---|---|---|---|---|---|---|
| Fitzgerald et al. 2006b (1Hz) | Magpro30 Medtronic | figure-eight coil | 1 | 110 | visual | DLPFC | Right | 5 cm | 20 | 1 | 900 | 1 | 0 | LF-rTMS-R |
| Fitzgerald et al. 2006b (2Hz) | Magpro30 Medtronic | figure-eight coil | 2 | 110 | visual | DLPFC | Right | 5 cm | 20 | 1 | 1800 | 1 | 0 | LF-rTMS-R |
| McDonald et al. 2006 | Neuronetics High Speed | figure-eight coil | L -10 R -1 | 110 | visual | DLPFC | BL | 5 cm | 10 | 1 | 1600 | | | LF-rTMS-R followed by HF-rTMS-L; HF-rTMS-L followed by LF-rTMS-R |
| Herwig et al. 2007 | Multiple | figure-eight coil | 10 | 110 | visual | DLPFC | Left | Beam F3 | 15 | 1 | 2000 | 100 | 8 | HF-rTMS-L |
| Dell Osso et al. 2009 | | figure-eight coil | 1 | 110 | EMG | DLPFC | Right | NN | 15 | 1 | 300 | 5 | 60 | LF-rTMS-R |
| Pallière Martinot et al. 2010 | MagStim Super Rapid | figure-eight coil | 10 | 90 | | DLPFC | | 5 cm | 10 | 1 | 1600 | | 60 | HF-rTMS |
| Harel et al. 2011 | MagStim Super Rapid 2 | H1 coil | 20 | 120 | EMG | DLPFC | Left | 5.5 cm | 20 | 1 | 1680 | 42 | 20 | HF-rTMS-L |
| Hernández-Ribas el al. 2013 | MagStim Super Rapid | figure-eight coil | 15 | 100 | visual | DLPFC | Left | 5 cm | 15 | 1 | 1500 | 20 | 60 | HF-rTMS-L |
| Ning et al. 2013 | MagStim Super Rapid 2 | figure-eight coil | 1 | 80 | | DLPFC | Right | | 20 | 1 | 1500 | 150 | 2 | LF-rTMS-R |

| | | | | | | | | | | | | | | |
|---|---|---|---|---|---|---|---|---|---|---|---|---|---|---|
| Beynel et al. 2014 | MagProX100,<br>MagVenture | figure-eight<br>coil | 50 | 80 | EMG | DLPFC | Left | NN | | 2 | 990 | 2 | 8 | iTBS-L |
| Speer et al. 2014 (L-HF) | Cadwell High Speed | figure-eight<br>coil | 20 | 110 | visual | DLPFC | Left | 5 cm | 15 | 1 | 1600 | 40 | 28 | HF-rTMS-<br>L |
| Chistyakov et al. 2015 | MagStim Super<br>Rapid 2 | figure-eight<br>coil | 50 | 100 | EMG | DLPFC | Right | 5 cm | 10 | 1 | 3600 | 4 | 900 | cTBS-R |
| Prasser et al. 2015 (LF/HF) | MagProX100,<br>MagVenture | figure-eight<br>coil | 1Hz/10Hz | 110 | EMG | DLPFC | BL | 6 cm | 15 | 1 | 2000 | | | F-rTMS-R<br>Followed<br>by HF-<br>rTMS-L |
| Prasser et al. 2015<br>(cTBS/iTBS) | MagProX100,<br>MagVenture | figure-eight<br>coil | | 80 | EMG | DLPFC | BL | 6 cm | 15 | 1 | 2400 | | | cTBS-R<br>Followed<br>by iTBS-L |
| Carnell et al. 2016 | | | R - 1<br>L - 1<br>BL - 10, 1 | 110 | visual | DLPFC | Right<br>Left<br>BL | 6 cm | 18 | BL - 2400 | BL - 2400 | R -1<br>L- 1<br>BL - 30,<br>1 | R -0<br>L-0<br>BL -<br>25, 0 | LF-rTMS-<br>R; LF-<br>rTMS-L;<br>HF-rTMS-<br>L<br>followed<br>by LF-<br>rTMS-R |
| Fitzgerald et al. 2016 | MagPro R30<br>MagVenture | figure-eight<br>coil | R – 1<br>L - 10 | 110 | | DLPFC | BL | NN | 20 | 2 | 2000 | R – 1<br>L- 20 | R- 0<br>L - 25 | LF-rTMS-<br>R<br>followed<br>by HF-<br>rTMS-L |
| Hu et al. 2016 (L-HF) | MagStim Rapid | figure-eight<br>coil | 10 | 80 | Visual | DLPFC | Left | 5.5cm | 20 | 1 | 1200 | 30 | 12 | HF-rTMS-<br>L |

| | | | | | | | | | | | | | | |
|---|---|---|---|---|---|---|---|---|---|---|---|---|---|---|
| Hu et al. 2016 (R-LF) | MagStim Rapid | figure-eight coil | 1 | 80 | visual | DLPFC | Right | 5.5cm | 20 | 1 | | 120 | 2 | LF-rTMS-R |
| Kazemi et al. 2016 (R-LF & L-HF) | MagStim Rapid | figure-eight coil | R – 1<br>L – 10 | R – 120<br>L – 100 | visual | DLPFC | BL | 5cm | 20 | 1 | 5250 | R – 150<br>L -75 | R – 2<br>L – 10 | LF-rTMS-R Followed by HF-rTMS-L |
| Kazemi et al. 2016 (R-LF) | MagStim Rapid | figure-eight coil | R - 1 | R - 120 | visual | DLPFC | Right | 5cm | 20 | 1 | 2500 | 250 | 2 | LF-rTMS-R |
| Rostami et al. 2017 | Neuro MS | figure-eight coil | R – 1<br>L – 10<br>Bilateral – 1, 10 | R – 120<br>L – 100<br>Bilateral – 100, 120 | visual | DLPFC | Right<br>Left<br>BL | 5cm | 20 | 1 | R – 2000<br>L – 3750<br>Bilateral - 5250 | R – 200<br>L – 75<br>Bilateral –R 150, L 75 | R -2<br>L – 15<br>Bilateral – R 2, L 15 | LF-rTMS-R; HF-rTMS-L; LF-rTMS-R Followed by HF-rTMS-L |
| Tavares et al. 2017 | Brainsway | H1 coil | 18 | 120 | visual | DLPFC | Left | 6cm | 20 | 1 | 1980 | 55 | 20 | HF-rTMS-L |
| Debaunes Jodoin et al. 2018 | MagProX100, MagVenture | figure-eight coil | 20 | 110 | visual | DLPFC | Left | BeamF3 | 20-30 | 2 | 3000 | 30 | 25 | HF-rTMS-L |
| Kazemi et al. 2018 | MagStim Rapid | figure-eight coil | R – 1<br>L - 10 | R – 120<br>L - 100 | visual | DLPFC | BL | F3 e F4 (EEG 10-20 system) | 10 | 1 | 5250 | R – 150<br>L - 75 | R – 2<br>L - 10 | LF-rTMS-R Followed by HF-rTMS-L |
| Rapinesi et al. 2018 | Brainsway | H1 coil | 20 | 120 | EMG | DLPFC | BL | 5.5cm | 20 | 1 | 2200 | 55 | 20 | HF-rTMS-BL |

| | | | | | | | | | | | | | | |
|---|---|---|---|---|---|---|---|---|---|---|---|---|---|---|
| Bulteau et al. 2019 | MagProX100, MagVenture | figure-eight coil | 50 | 80 | | DLPFC | Left | NN | 30 | 2 | 990 | | 10 | iTBS-L |
| Kito et al. 2019 (ITI=11s) | MagPro R30, MagVenture | figure-eight coil | 10 | 120 | | DLPFC | Left | 5cm | 20-30 | 1 | 3000 | 75 | 11 | HF-rTMS-L |
| Kito et al. 2019 (ITI=26s) | MagPro R30, MagVenture | figure-eight coil | 10 | 120 | | DLPFC | Left | 5cm | 20-30 | 1 | 3000 | 75 | 26 | HF-rTMS-L |
| Goldwaser et al. 2020 | Neuronetics Neurostar | figure-eight coil | 10 | 120 | | DLPFC | Left | BeamF3 | 30 | 1 | 3000 | 75 | 26 | HF-rTMS-L |
| Olejarczyk et al. 2020 | MagProX100, MagVenture | | 10 | 120 | | DLPFC | Left | | 20 | 1 | 2000 | 1 | 0 | HF-rTMS-L |
| Phillips et al. 2020 | Neuronetics Neurostar | | 10 | 100<br>120 | Visual | DLPFC | Left | BeamF3 | 30 | 1 | | | | HF-rTMS-L |
| Mak et al. 2021 | MagStim Super Rapid 2 | figure-eight coil | 1 | 110 | EMG | DLPFC | Right | NN | 15 | 1 | 300 | 5 | 60 | LF-rTMS-R |
| McGirr et al. 2021 | MagProX100, MagVenture | figure-eight coil | 50 | 120 | EMG & visual | DLPFC | Left | NN | 20 | 1 | 600 | | 8 | iTBS-L |
| Yang et al. 2021 | MagStim Super Rapid 2 | | 10 | 120 | | DLPFC | Left | 6cm | | 1 | 3000 | 75 | 26 | HF-rTMS-L |
| Abdullah Alhelali et al. 2022 | | | | | | | | | 19 | | 1935 | | | |
| Gama-Chonlon et al. 2022 (10 Hz) | | figure-eight coil | 10 | 120 | | DLPFC | Left | Beam F3 | 30 | | 3000 | | | HF-rTMS-L |
| Gama-Chonlon et al. 2022 (R-LF & L-HF) | | figure-eight coil | R – 1<br>L - 10 | R – 110<br>L - 120 | | DLPFC | BL | | 30 | | 3900 | | | |

| | | | | | | | | | | | | | | |
|---|---|---|---|---|---|---|---|---|---|---|---|---|---|---|
| Gama-Chonlon et al. 2022 (mixed) | | figure-eight coil | | | | | | | 30 | | | | | |
| Koutsomitros et al. 2022 | MagPro R20 MagVenture MagPro R30 MagVenture | figure-eight coil | 10 | 120 | | DLPFC | Left | F3 e F4 (EEG 10-20 system) | 20 | 1 | 1000 | 20 | 11 | HF-rTMS-L |
| Zengin et al. 2022 | Neuro MS/D | figure-eight coil | 10 | 110 | visual | DLPFC | Left | 5cm | 20 | 2 | 1000 | 25 | 20 | HF-rTMS-L |
| Bouaziz et al. 2023 (1Hz) | MagProX100, MagVenture | | 1 | 120 | Visual | DLPFC | Right | 5/6 cm | | 1 | 360 | | | LF- rTMS -R |
| Bouaziz et al. 2023 (10Hz) | MagProX100, MagVenture MagStim Super Rapid | | 10 | 110-120 | Visual | DLPFC | Left | 5/6 cm or NN | | 1 | 1600-2000 | | | HF-rTMS-L |
| Bouaziz et al. 2023 (20Hz) | MagProX100, MagVenture | | 20 | 90 | Visual | DLPFC | Left | 5/6 cm or NN | | 1 | 800 | 20 | 60 | HF-rTMS-L |
| Bouaziz et al. 2023 (iTBS) | MagProX100, MagVenture MagStim Super Rapid | | 50 | 80 | visual | DLPFC | Left | 5/6 cm or NN | | 1 | 600-1620 | | | HF-rTMS-L |
| Mallik et al. 2023 | MagPro R30, MagVenture | figure-eight coil | 50 | 80 | EMG | DLPFC | Right | F3 e F4 (EEG 10-20 system) | 15 | 3 | 1800 | | | cTBS-R |
| Aaronson et al. 2024 | Neuronetics Neurostar | figure-eight coil | 10 | 120 | visual | DLPFC | Left | 5,5 cm | 35 | 1 | 3000 | 75 | 26 | HF-rTMS-L |

| | | | | | | | | | | | | | | |
|---|---|---|---|---|---|---|---|---|---|---|---|---|---|---|
| Ikawa et al. 2024 | MagProX100, MagVenture | figure-eight coil | 1 | 120 | | DLPFC | Right | Beam F3 inverted | 30 | 1 | 600 | | | LF-rTMS-R |
| Dellink et al. 2024 | Magstim Super Rapid 2 Plus | figure-eight coil | 50 | 110 | visual | DLPFC | Right | NN | 20 | 5 | 900 | 1 | | cTBS-R |
| Novak et al. 2024 (RVL) | MagPro R30 MagVenture | figure-eight coil | 10 | 100 | EMG | RVLPFC | Right | NN | 20 | 1 | 1200 | | 8 | HF-rTMS-R |
| Novak et al. 2024 (LDL) | MagPro R30 MagVenture | figure-eight coil | 10 | 100 | EMG | DLPFC | Left | NN | 20 | 1 | 1200 | | 8 | HF-rTMS-L |
| Sheline et al. 2024 | MagProX100, MagVenture | figure-eight coil | 50 | 90 | | DLPFC | Left | NN | 50 | 10 | 1800 | | | iTBS-L |

BL – Bilateral; cTBS – continuous Theta Burst Stimulation; DLPFC – Dorsolateral prefrontal cortex; EMG – electromyography; HF – High frequency; iTBS – intermittent Theta Burst Stimulation; ITI - Inter-train interval; L – Left; LDL – left dorsolateral; LF – Low frequency; NN – NeuroNavegationNeuronavigation; No. – Number; R – Right; rMT – resting Motor Threshold; rTMS – repetitive Transcranial Magnetic Stimulation; RVL – right ventrolateral; Stim – Stimulation.

**Table S6** – Study quality assessment by Cochrane risk-of-bias tool for randomized trials (RoB2).

| Authors, year | Domain 1 | Domain 2 | Domain 3 | Domain 4 | Domain 5 | Overall risk of bias |
|---|---|---|---|---|---|---|
| Kimbrell et al. 1999 | | | | | | |
| Klein et al. 1999 | | | | | | |
| George et al. 2000 | | | | | | |
| Dolberg et al. 2000 | | | | | | |
| Loo et al. 2003 | | | | | | |
| Nahas et al. 2003 | | | | | | |
| Rossini et al. 2005 | | | | | | |
| Su et al. 2005 | | | | | | |
| Fitzgerald et al. 2006a | | | | | | |
| Fitzgerald et al. 2006b | | | | | | |
| McDonald et al. 2006 | | | | | | |
| Herwig et al. 2007 | | | | | | |
| Pallière Martinot et al. 2010 | | | | | | |
| Hernandez Ribas el al. 2013 | | | | | | |
| Ning et al. 2013 | | | | | | |
| Speer et al. 2014 | | | | | | |
| Chistyakov et al. 2015 | | | | | | |
| Prasser et al. 2015 | | | | | | |
| Beynel et al. 2014 | | | | | | |
| Fitzgerald et al. 2016 | | | | | | |
| Hu et al. 2016 | | | | | | |
| Kazemi et al. 2016 | | | | | | |

Tavares et al. 2017
Bulteau et al. 2019
Kito et al. 2019
Mak et al. 2021
McGirr et al. 2021
Zengin et al. 2022
Mallik et al. 2023
Dellink et al. 2024
Novak et al. 2024
Sheline et al. 2024

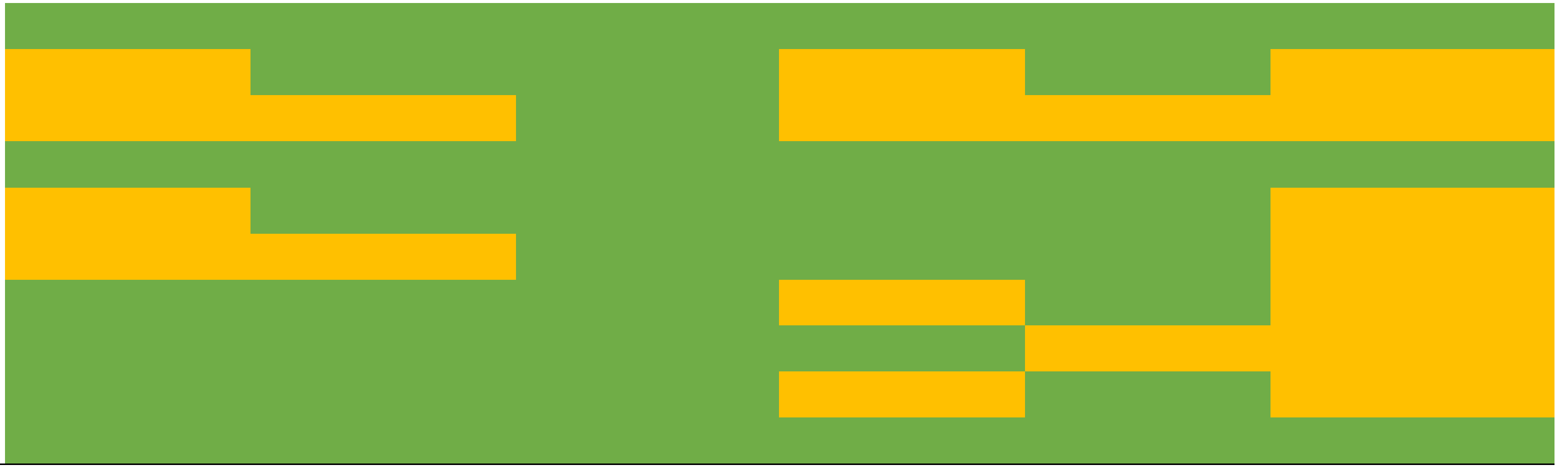

| Overall risk-of-bias judgement | Criteria |
|---|---|
| **Low risk of bias** | The study is judged to be at low risk of bias for all domains for this result. |
| **Some concerns** | The study is judged to raise some concerns in at least one domain for this result, but not to be at high risk of bias for any domain. |
| **High risk of bias** | The study is judged to be at high risk of bias in at least one domain for this result.<br>Or<br>The study is judged to have some concerns for multiple domains in a way that substantially lowers confidence in the result. |

**Table S7** – Study quality assessment by Newcastle-Ottawa Quality Assessment Scale for cohort studies

| Authors, year | Selection | | | | | | | | | | | | | | | | | | Comparability | | | | Outcome | | | | | | | | | | | | | | Total* (0-9) |
|---|---|---|---|---|---|---|---|---|---|---|---|---|---|---|---|---|---|---|---|---|---|---|---|---|---|---|---|---|---|---|---|---|---|---|---|---|---|
| | 1 | | | | | 2 | | | | 3 | | | | | 4 | | | Total* (0-4) | 1 | | | Total* (0-2) | 1 | | | | | 2 | | | 3 | | | | | Total* (0-3) | |
| | a) | b) | c) | d) | * | a) | b) | c) | * | a) | b) | c) | d) | * | a) | b) | * | | a) | b) | * | | a) | b) | c) | d) | * | a) | b) | * | a) | b) | c) | d) | * | | |
| Dell Osso et al. 2009 | - | 1 | - | | 1 | - | - | - | 0 | - | 1 | - | - | 1 | 1 | - | 1 | 3 | - | - | 0 | 0 | - | - | 1 | - | 0 | 1 | - | 1 | - | - | - | 1 | 0 | 1 | 4 |
| Harel et al. 2011 | - | 1 | - | - | 1 | 1 | - | - | 1 | - | 1 | - | - | 1 | 1 | - | 1 | 4 | 1 | 1 | - | 2 | - | - | 1 | - | 0 | 1 | - | 1 | - | 1 | - | - | 1 | 2 | 8 |
| Carnell et al. 2016 | - | 1 | - | | 1 | - | - | - | 0 | - | 1 | - | - | 1 | 1 | - | 1 | 3 | - | - | 0 | 0 | - | - | 1 | - | 0 | 1 | - | 1 | - | - | - | 1 | 0 | 1 | 4 |
| Rostami et al. 2017 | 1 | - | - | - | 1 | - | - | - | 0 | 1 | - | - | - | 1 | 1 | - | 1 | 3 | - | - | 0 | 0 | - | - | 1 | - | 0 | 1 | - | 1 | - | 1 | - | - | 1 | 2 | 5 |
| Debaunes Jodoin et al. 2018 | - | 1 | - | - | 1 | - | - | - | 0 | - | 1 | - | - | 1 | 1 | - | 1 | 3 | - | 1 | 1 | 1 | - | - | 1 | - | 0 | 1 | - | 1 | 1 | - | - | - | 1 | 2 | 6 |
| Kazemi et al. 2018 | - | 1 | - | - | 1 | - | 1 | - | 0 | - | 1 | - | - | 1 | - | 1 | 0 | 2 | - | 1 | 1 | 1 | - | - | 1 | - | 0 | 1 | - | 1 | - | 1 | - | - | 1 | 2 | 5 |
| Rapinesi et al. 2018 | - | 1 | - | - | 1 | - | - | - | 0 | - | 1 | - | - | 1 | 1 | - | 1 | 3 | - | - | 0 | 0 | - | - | 1 | - | 0 | 1 | - | 1 | 1 | - | - | - | 1 | 2 | 5 |
| Goldwaser et al. 2020 | - | 1 | - | - | 1 | - | - | - | 0 | - | - | - | 1 | 0 | 1 | - | 1 | 2 | - | - | 0 | 0 | - | 1 | - | - | 1 | 1 | - | 1 | - | 1 | - | - | 1 | 3 | 5 |
| Olejarczyk et al. 2020 | - | 1 | - | - | 1 | - | - | - | 0 | 1 | - | - | - | 1 | - | 1 | 0 | 2 | - | - | 0 | 0 | - | - | 1 | - | 0 | - | 1 | 0 | - | 1 | - | - | 1 | 1 | 3 |

| | | | | | | | | | | | | | | | | | | | | | | | | | | | | | | | | | | | | | |
|---|---|---|---|---|---|---|---|---|---|---|---|---|---|---|---|---|---|---|---|---|---|---|---|---|---|---|---|---|---|---|---|---|---|---|---|---|---|
| Phillips et al. 2020 | - | 1 | - | - | 1 | - | - | - | 0 | - | - | - | 1 | 0 | 1 | - | 1 | 2 | - | 1 | 1 | 1 | - | - | 1 | - | 0 | 1 | - | 1 | - | - | - | 1 | 0 | 1 | 4 |
| Yang et al. 2021 | - | 1 | - | - | 1 | - | - | - | 0 | - | - | - | 1 | 0 | - | 1 | 0 | 1 | - | 1 | 1 | 1 | - | - | 1 | - | 0 | 1 | - | 1 | - | 1 | - | - | 1 | 2 | 4 |
| Abdullah Alhelali et al. 2022 | - | 1 | - | - | 1 | - | - | - | 0 | 1 | - | - | - | 1 | - | 1 | 0 | 2 | - | - | 0 | 0 | - | 1 | - | - | 1 | 1 | - | 1 | 1 | - | - | - | 1 | 3 | 5 |
| Gama-Chonlon et al. 2022 | - | 1 | - | - | 1 | - | - | 1 | - | - | - | 1 | - | 0 | 1 | - | 1 | 2 | 1 | 1 | 2 | 2 | - | - | - | 1 | 0 | 1 | - | 1 | - | 1 | - | - | 1 | 2 | 6 |
| Koutsomitros et al. 2022 | - | 1 | - | - | 1 | - | - | 0 | 0 | 1 | - | - | - | 1 | 1 | - | 1 | 4 | 1 | 1 | 2 | 2 | 1 | - | - | - | 1 | 1 | - | 1 | 1 | - | - | - | 1 | 2 | 8 |
| Bouaziz et al. 2023 | - | 1 | - | - | 1 | - | - | 1 | 0 | 1 | - | - | - | 1 | 1 | - | 1 | 3 | 1 | 1 | 2 | 2 | - | 1 | - | - | - | - | - | 0 | - | - | 1 | 0 | 1 | 1 | 6 |
| Aaronson et al. 2024 | 1 | - | - | - | 1 | - | - | - | 0 | - | 1 | - | - | 1 | 1 | - | 1 | 3 | 1 | 1 | 2 | 2 | - | - | - | 1 | 0 | 1 | - | 1 | - | 1 | - | - | 1 | 2 | 7 |
| Ikawa et al. 2024 | - | 1 | - | - | 1 | 1 | - | - | 1 | 1 | - | - | - | 1 | 1 | - | 1 | 4 | - | - | 0 | 0 | 1 | - | - | - | 1 | - | 1 | 0 | - | - | - | 1 | 0 | 1 | 5 |

| Overall risk-of-bias judgement | Scores |
|---|---|
| **Low quality and high risk of bias** | 0-3 |
| **Moderate quality and moderate risk of bias** | 4-6 |
| **High quality and low risk of bias** | 7-9 |

**Selection**

1) Is the case definition adequate?

a) yes, with independent validation **

b) yes, eg record linkage or based on self-reports

c) no description

2) Representativeness of the cases

a) consecutive or obviously representative series of cases **

b) potential for selection biases or not stated

3) Selection of Controls

a) community controls **

b) hospital controls

c) no description

4) Definition of Controls

a) no history of disease (endpoint) **

b) no description of source

**Comparability**

1) Comparability of cases and controls on the basis of the design or analysis

a) study controls for _______________ (Select the most important factor.) **

b) study controls for any additional factor ** (This criteria could be modified to indicate specific control for a second important factor.)

**Exposure**

1) Ascertainment of exposure

a) secure record (eg surgical records) **

b) structured interview where blind to case/control status **

c) interview not blinded to case/control status

d) written self report or medical record only

e) no description

2) Same method of ascertainment for cases and controls

a) yes **

b) no

3) Non-Response rate

a) same rate for both groups **

b) non respondents described

c) rate different and no designation

**Table S8 -** Meta-regression for dichotomous variables.

| Dichotomous variables* | Response rate | | | Remission rate | | | Dep. improvement | | |
|---|---|---|---|---|---|---|---|---|---|
| | β±SE | p | N | β±SE | p | N | β±SE | p | N |
| **Clinical** | | | | | | | | | |
| TRD inclusion (Yes vs. No) | -6.84±8.97 | 0.45 | 46 | -7.27±7.05 | 0.31 | 32 | -0.15±0.37 | 0.68 | 33 |
| **Transcranial Magnetic Stimulation** | | | | | | | | | |
| MT Determination (EMG vs. Visual) | -10.93±9.78 | 0.27 | 39 | -9.40±8.75 | 0.29 | 26 | 0.46±0.30 | 0.14 | 26 |
| Accelerated Protocol (Yes vs. No) | -5.34±11.27 | 0.64 | 54 | -9.44±10.32 | 0.37 | 37 | -0.29±0.41 | 0.48 | 35 |
| Use of Neuronavigation (Yes vs. No) | -14.27±10.06 | 0.16 | 48 | -15.10±8.25 | 0.08 | 31 | 0.15±0.35 | 0.66 | 28 |
| Frequency (HF vs. LF)** | -13.69±11.23 | 0.23 | 40 | -6.87±10.48 | 0.52 | 31 | -0.96±0.50 | 0.07 | 28 |
| Laterality (BL vs. UL) | -13.22±11.02 | 0.24 | 53 | -19.35±10.41 | 0.07 | 37 | 0.29±0.56 | 0.61 | 34 |
| Side (Left vs. Right) | -3.50±9.65 | 0.72 | 44 | 0.57±9.52 | 0.95 | 32 | -0.57±0.41 | 0.17 | 30 |
| **Study Characteristics** | | | | | | | | | |
| Study Type (RCT vs. Non-RCT) | -7.20±7.80 | 0.36 | 58 | -9.83±7.27 | 0.18 | 40 | 0.28±0.31 | 0.38 | 39 |

BL – Bilateral; EMG – Electromyography; HF – High Frequency; LF – Low Frequency; RCT – Randomized Clinical Trial; TRD - Treatment resistant depression; UL - Unilateral

*each variable was coded as 1 and 0; coefficients represent the effect of being in level 1 (underlined) compared to level 0.

**variable was categorized into 2 levels (low frequency: <5Hz; high frequency: >5Hz); a single 5Hz study (N=1) was excluded from frequency analysis since there is no clear consensus on frequency level classification and modulatory effects of such protocols (21,22)

### Supplementary Figures

**Figure S1 – Publication bias assessment.** Publication bias can be assessed using visual inspection of funnel plots when an adequate number of studies (N≥10) are available (20).

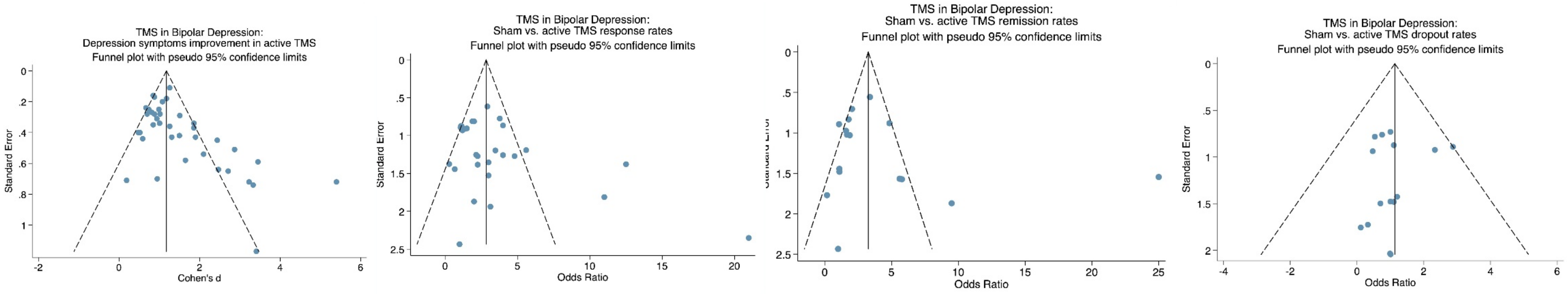

**Figure S2 - Forest plot of random effects meta-analyses.** Treatment emergent mania rates were similar between active and sham.

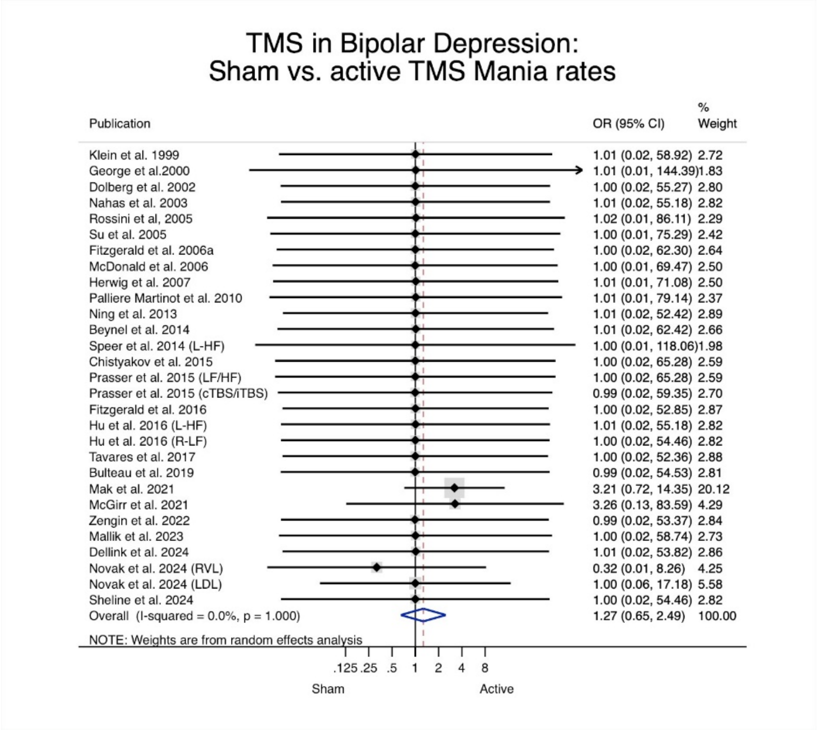

### Supplementary References


1. McGirr A, Vila-Rodriguez F, Cole J, Torres IJ, Arumugham SS, Keramatian K, et al. Efficacy of Active vs Sham Intermittent Theta Burst Transcranial Magnetic Stimulation for Patients With Bipolar Depression: A Randomized Clinical Trial. JAMA Netw Open. 2021;4(3):e210963.
2. Yang YB, Chan P, Rayani K, McGirr A. Comparative Effectiveness of Repetitive Transcranial Magnetic Stimulation in Unipolar and Bipolar Depression. Can J Psychiatry. 66. United States2021. p. 313-5.
3. Kimbrell TA, Little JT, Dunn RT, Frye MA, Greenberg BD, Wassermann EM, et al. Frequency dependence of antidepressant response to left prefrontal repetitive transcranial magnetic stimulation (rTMS) as a function of baseline cerebral glucose metabolism. Biol Psychiatry. 1999;46(12):1603-13.
4. Klein E, Kreinin I, Chistyakov A, Koren D, Mecz L, Marmur S, et al. Therapeutic efficacy of right prefrontal slow repetitive transcranial magnetic stimulation in major depression: a double-blind controlled study. Arch Gen Psychiatry. 1999;56(4):315-20.
5. George MS, Nahas Z, Molloy M, Speer AM, Oliver NC, Li XB, et al. A controlled trial of daily left prefrontal cortex TMS for treating depression. Biol Psychiatry. 2000;48(10):962-70.
6. Loo CK, Mitchell PB, Croker VM, Malhi GS, Wen W, Gandevia SC, et al. Double-blind controlled investigation of bilateral prefrontal transcranial magnetic stimulation for the treatment of resistant major depression. Psychol Med. 2003;33(1):33-40.
7. Rossini D, Lucca A, Zanardi R, Magri L, Smeraldi E. Transcranial magnetic stimulation in treatment-resistant depressed patients: a double-blind, placebo-controlled trial. Psychiatry Res. 2005;137(1-2):1-10.
8. Su TP, Huang CC, Wei IH. Add-on rTMS for medication-resistant depression: a randomized, double-blind, sham-controlled trial in Chinese patients. J Clin Psychiatry. 2005;66(7):930-7.
9. McDonald WM, Easley K, Byrd EH, Holtzheimer P, Tuohy S, Woodard JL, et al. Combination rapid transcranial magnetic stimulation in treatment refractory depression. Neuropsychiatr Dis Treat. 2006;2(1):85-94.
10. Paillère Martinot ML, Galinowski A, Ringuenet D, Gallarda T, Lefaucheur JP, Bellivier F, et al. Influence of prefrontal target region on the efficacy of repetitive transcranial magnetic stimulation in patients with medication-resistant depression: a [(18)F]-fluorodeoxyglucose PET and MRI study. Int J Neuropsychopharmacol. 2010;13(1):45-59.
11. Hernández-Ribas R, Deus J, Pujol J, Segalàs C, Vallejo J, Menchón JM, et al. Identifying brain imaging correlates of clinical response to repetitive transcranial magnetic stimulation (rTMS) in major depression. Brain Stimul. 2013;6(1):54-61.
12. Chistyakov AV, Kreinin B, Marmor S, Kaplan B, Khatib A, Darawsheh N, et al. Preliminary assessment of the therapeutic efficacy of continuous theta-burst magnetic stimulation (cTBS) in major depression: a double-blind sham-controlled study. J Affect Disord. 2015;170:225-9.

13. Prasser J, Schecklmann M, Poeppl TB, Frank E, Kreuzer PM, Hajak G, et al. Bilateral prefrontal rTMS and theta burst TMS as an add-on treatment for depression: a randomized placebo controlled trial. World J Biol Psychiatry. 2015;16(1):57-65.
14. Bulteau S, Beynel L, Marendaz C, Dall'Igna G, Peré M, Harquel S, et al. Twice-daily neuronavigated intermittent theta burst stimulation for bipolar depression: A Randomized Sham-Controlled Pilot Study. Neurophysiol Clin. 2019;49(5):371-5.
15. Herwig U, Fallgatter AJ, Höppner J, Eschweiler GW, Kron M, Hajak G, et al. Antidepressant effects of augmentative transcranial magnetic stimulation: randomised multicentre trial. Br J Psychiatry. 2007;191:441-8.
16. Desbeaumes Jodoin V, Miron JP, Lespérance P. Safety and Efficacy of Accelerated Repetitive Transcranial Magnetic Stimulation Protocol in Elderly Depressed Unipolar and Bipolar Patients. Am J Geriatr Psychiatry. 2019;27(5):548-58.
17. Kito S, Miyazi M, Nakatani H, Matsuda Y, Yamazaki R, Okamoto T, et al. Effectiveness of high-frequency left prefrontal repetitive transcranial magnetic stimulation in patients with treatment-resistant depression: A randomized clinical trial of 37.5-minute vs 18.75-minute protocol. Neuropsychopharmacol Rep. 2019;39(3):203-8.
18. Bouaziz N, Laidi C, Bulteau S, Berjamin C, Thoms F, Moulier V, et al. Real world transcranial magnetic stimulation for major depression: a multisite naturalistic, retrospective study. Brain Stimulation. 2023;16(1):384-5.
19. Dellink A, Hebbrecht K, Zeeuws D, Baeken C, De Fré G, Bervoets C, et al. Continuous theta burst stimulation for bipolar depression: A multicenter, double-blind randomized controlled study exploring treatment efficacy and predictive potential of kynurenine metabolites. J Affect Disord. 2024;361:693-701.
20. Dalton JE, Bolen SD, Mascha EJ. Publication Bias: The Elephant in the Review. Anesth Analg. 123. United States2016. p. 812-3.
21. Di Lazzaro V, Dileone M, Pilato F, Capone F, Musumeci G, Ranieri F, et al. Modulation of motor cortex neuronal networks by rTMS: comparison of local and remote effects of six different protocols of stimulation. J Neurophysiol. 2011;105(5):2150-6.
22. Fitzgerald PB, Fountain S, Daskalakis ZJ. A comprehensive review of the effects of rTMS on motor cortical excitability and inhibition. Clin Neurophysiol. 2006;117(12):2584-96.
